# A Data-Driven Biopsychosocial Framework Determining the Spreading of Chronic Pain

**DOI:** 10.1101/2022.07.22.22277850

**Authors:** Christophe Tanguay-Sabourin, Matt Fillingim, Marc Parisien, Gianluca V Guglietti, Azin Zare, Jax Norman, Ronrick Da-ano, Jordi Perez, Scott J Thompson, Marc O Martel, Mathieu Roy, Luda Diatchenko, Etienne Vachon-Presseau

## Abstract

Chronic pain conditions are complex syndromes characterized by a mosaic of biological, psychological, and social factors. We derived predictive models for the number of co- existing pain sites in the UK Biobank and identified a common risk score that classified different chronic pain conditions in cross-sectional data, predicted the development of chronic pain in pain-free individuals, and determined the spreading of chronic pain to multiple sites or its recovery nine years later. The features with the strongest prognosis included sleeplessness, feeling ‘fed-up’, tiredness, stressful life events, and a BMI > 30. The risk score for pain was associated with an inflammatory blood marker, a polygenic risk score for pain, and a neuroimaging-based marker for sustained pain. The demonstration of a common biopsychosocial risk factor for different clinical pain conditions may help better characterize a general chronic pain syndrome, tailor research protocols, optimize patient randomization in clinical trials, and improve pain management.

---

Pain is the primary reason individuals seek healthcare and the most important source of disability among working adults ^1–3^. Unfortunately, the cause of chronic pain and its prognosis often remains unknown, as tissue damage following injury is a poor predictor of clinical outcomes ^4^. The interactions between biological, psychological, and social factors instead better determine chronic pain conditions and the functioning of the patients ^5^. This holistic framework is often referred to as the biopsychosocial model for chronic pain ^6^. Despite its popularity, the elements composing the model remain hard to define due to the difficulties of simultaneously measuring and dissociating multidimensional factors in large groups of pain patients. Recent accessibility to large cohorts of chronic pain patients provides unprecedented opportunities to tackle these problems and better understand the determinants of chronic pain.

Prognosis studies have shown that maladaptive pain coping strategies, somatization of pain, and history of pain increases the likelihood of developing long term back pain ^4^. For instance, pain severity and duration ^7–10^, fear of pain ^11^, and pain catastrophizing ^4, 12^ have all been associated with worsening of back pain. Moreover, brain imaging and genetic studies have also suggested that biological factors predispose to chronic pain conditions ^13^. Yet, these studies are often circular, as the features entered to predict pain are mostly pain measurements or attitudes towards pain, and most of the candidate brain imaging markers for chronic pain have been identified in small sample studies that hinders their reproducibility in larger heterogenous sample of participants ^14, 15^. Moreover, these previous prospective studies have rarely been validated in out-of-sample patients and their generalization to new patients remains unknown. Large scale longitudinal studies examining the contribution of candidate biological markers to the development of chronic pain and their associations with psychosocial factors predictive of pain are currently lacking.

The Task Force for the Classification of Chronic Pain recommended that chronic pain conditions be classified based on their etiology (*i.e.,* visceral pain), underlying pathophysiology (*i.e.,* neuropathic pain), or body site (*i.e.,* knee pain) ^16, 17^. Despite notable differences between these conditions, the evidence also suggests that different pain conditions are overlapping with one another ^19^, sharing a common genetic risk factor ^20, 21^, and showing similar alterations in the central nervous system ^13, 22, 23^. Thus, different conditions share common risk factors and primary chronic pain has been recognized as a disease on its own term rather than the symptom of another disease ^18^. Here, we used a data-driven approach that synthesized a wide range of multidimensional pain-agnostic features to derive empirical models for classifying and forecasting different pain conditions. The reproducibility and the generalizability of these predictive models were estimated in a large number of participants reporting pain and outline a robust roadmap characterizing a general *chronic pain syndrome*. We first hypothesized that different chronic pain conditions are characterized by common psychosocial factors that can be observed and identified by studying the number of co-existing pain sites. We secondly hypothesized that these psychosocial risk score can predict the development of different chronic pain conditions. The mismatch between the baseline pain levels and the individual psychosocial risk score will determine the spreading or the recovering of chronic pain across body sites in the longitudinal data. We finally hypothesized that biological markers for chronic pain are associated with the identified psychosocial features predictive of the different chronic pain conditions.

To test these hypotheses, we used machine learning to train a biopsychosocial model on the number of co-existing pain sites to derive a general risk score for pain. The risk score for pain could classify each chronic pain condition separately in cross-sectional data (seven different body sites and 25 pain-related ICD-10 diagnoses) and forecast individual differences in the spreading or recovery of chronic pain nine years later. Our model also generalized to secondary outcomes such as general health, disability, and opioid use. We then showed that our risk score for pain was associated with three pre-selected biological factors available in the biobank, namely, an inflammatory blood marker, a polygenic risk score computed for the number of pain sites, and a neuroimaging-based marker for sustained pain. We finally show that a sparse model based on six items (with binarized answers) can account for different pain conditions and predict the development of chronic pain in healthy individuals. Overall, our study provides a comprehensive biopsychosocial framework that characterizes a general *chronic pain syndrome* in a large and representative sample of patients in the UK Biobank.

## Results

This study was conducted using data from the UK Biobank (UKBB). On their initial visit, participants were asked if they experienced pain interfering with their usual activities at the following body sites: headache, face, neck/shoulder, stomach/abdominal, back, hip, and knee. The participants could also respond that they experienced pain all over the body or none of the above (these were categorized as pain-free participants). **Fig. 1a** shows the prevalence of pain in the full sample of participants (n = 493,211) and a subsample of participants that returned for a follow up magnetic resonance imaging (MRI) visit about nine years later (n = 48,079). Participants reporting pain were then asked if they had pain lasting for more than 3 months, which represents the cut-off for the pain to be considered *chronic* ^16^. Pain experienced for less than 3 months was therefore considered *acute*.

**Fig. 1.**
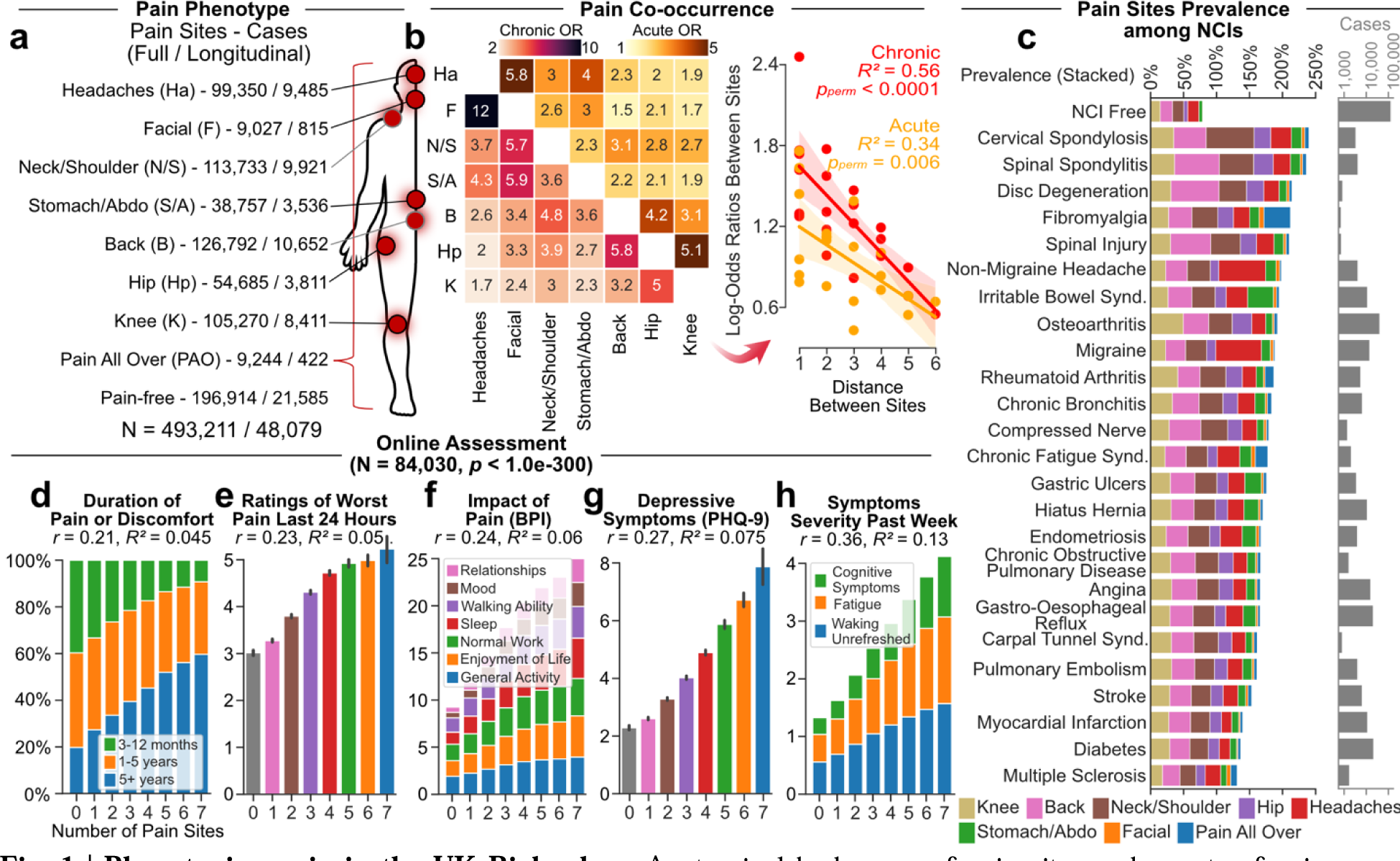
Phenotyping pain in the UK Biobank. **a.** Anatomical body map of pain sites and counts of pain cases (acute and chronic) for the full sample and for individuals with a follow-up visit nine years later. **b.** Odds ratios of co-occurrence between pain sites (chronic on the left and acute on right) at baseline. The log-odds ratios of co- occurring pain between two sites were negatively associated with their distances. Significance was determined using 10,000 permutation tests. **c.** The prevalence of pain is shown per body sites among 25 Non-Cancer Illnesses (NCI) commonly associated with chronic pain and the count of cases reported. **d-h.** In the online follow-up pain questionnaires, the number of co-existing pain sites was associated with **d.** the duration or discomfort of pain, **e.** ratings of the least pain on 10 in the last 24 hours **f.** the interference of pain across 7 dimensions, **g.** depressive symptoms severity in last 2 weeks, and **h.** symptoms severity during the last week. Abbreviations: Headache (Ha); Facial (F); Neck or Shoulder (N/S); Stomach or Abdominal (S/A); Back (B); Hip (Hp); Knee (K); Non-Cancer Illness (NCI); Brief Pain Inventory (BPI); Patient Health Questionnaire (PHQ).

### Multi-site pain in the UKBB

We first aimed to derive a model that was predictive of all chronic pain conditions classified from their body sites. To this end, we trained a model that predicted the number of co-existing pain sites reported by the patient to identify common features between co-existing pain sites. Previous studies have shown that over one third of chronic pain patients report co-existing pain conditions associated with lower quality of life and poorer response to treatment ^19, 24^. In the UK Biobank, 44% of chronic pain patients reported pain at more than one body site and the co-occurrence of pain was more frequent between proximal sites than distal sites (**fig. 1b,c**). These results emphasize that pain was not amplified uniformly across body sites, an effect that was also observed in acute pain conditions. We next examined the prevalence of these pain conditions across a series of clinical conditions diagnosed according to the international classification of diseases (ICD-10). Here, pain conditions and other pain-related non-cancer illnesses (NCI) were all characterized by overlapping pain conditions (**fig. 1c**). This can be appreciated in the case of migraine, non-migraine headache, or spinal spondylitis, where the prevalence of pain at the head (migraine, headache) or back (spinal spondylitis) sites was lower than the cumulative prevalence of pain at the remaining sites (**fig. 1c**).

The importance of co-existing pain conditions was then examined using the online follow up data collected about 12 years after the initial visit (n = 84,030 chronic pain patients, excluding pain all over the body). The number of pain sites reported at the time of the online follow-up questionnaires showed a monotonic increase with pain duration (**fig. 1d**), pain intensity (**fig. 1e**), impact of pain (**fig. 1f**), depressive symptoms (**fig. 1g**), and symptoms severity (**fig. 1h**). Higher-resolution anatomical body sites available in the online questionnaire further confirmed the spatial co-occurrence and interdependence in pain ratings across sites (**extended fig.1**). Here again, diagnosed clinical conditions such as pelvic pain or carpal tunnel syndrome were characterized by co-existing pain at other body sites. Overall, these results show that the number of co-existing pain sites is an important phenotype that characterizes different chronic pain conditions and reflects the severity and the impact of these pain conditions. We conclude that the number of co-existing pain site may therefore be used to derive our biopsychosocial framework characterizing a general *chronic pain syndrome*.

### Predicting the number of pain sites

We applied machine learning algorithms on 99 non-pain-related features assessing physical, psychological, demographic, and sociological factors to derive a risk score that predicted the number of pain sites. To this end, the UKBB dataset available at the baseline visit was divided into a training set (n = 445,132) for discovery and a testing set composed of out-of-sample participants for whom longitudinal data were available (n = 48,079). In the discovery set, we applied Nonlinear Iterative Partial Least Square (NIPALS) regression algorithm on the 99 features to predict the number of co-existing pain sites (combining acute and chronic). This was done using a 10-folds cross-validation within the training set to estimate the model fit and identify the optimal number of components to retain in an unbiased manner (**extended fig. 2**). The derived model applied in the test set accounted for about 14% of the variance in the number of co-existing pain sites (**fig. 2a-c**).

**Fig. 2.**
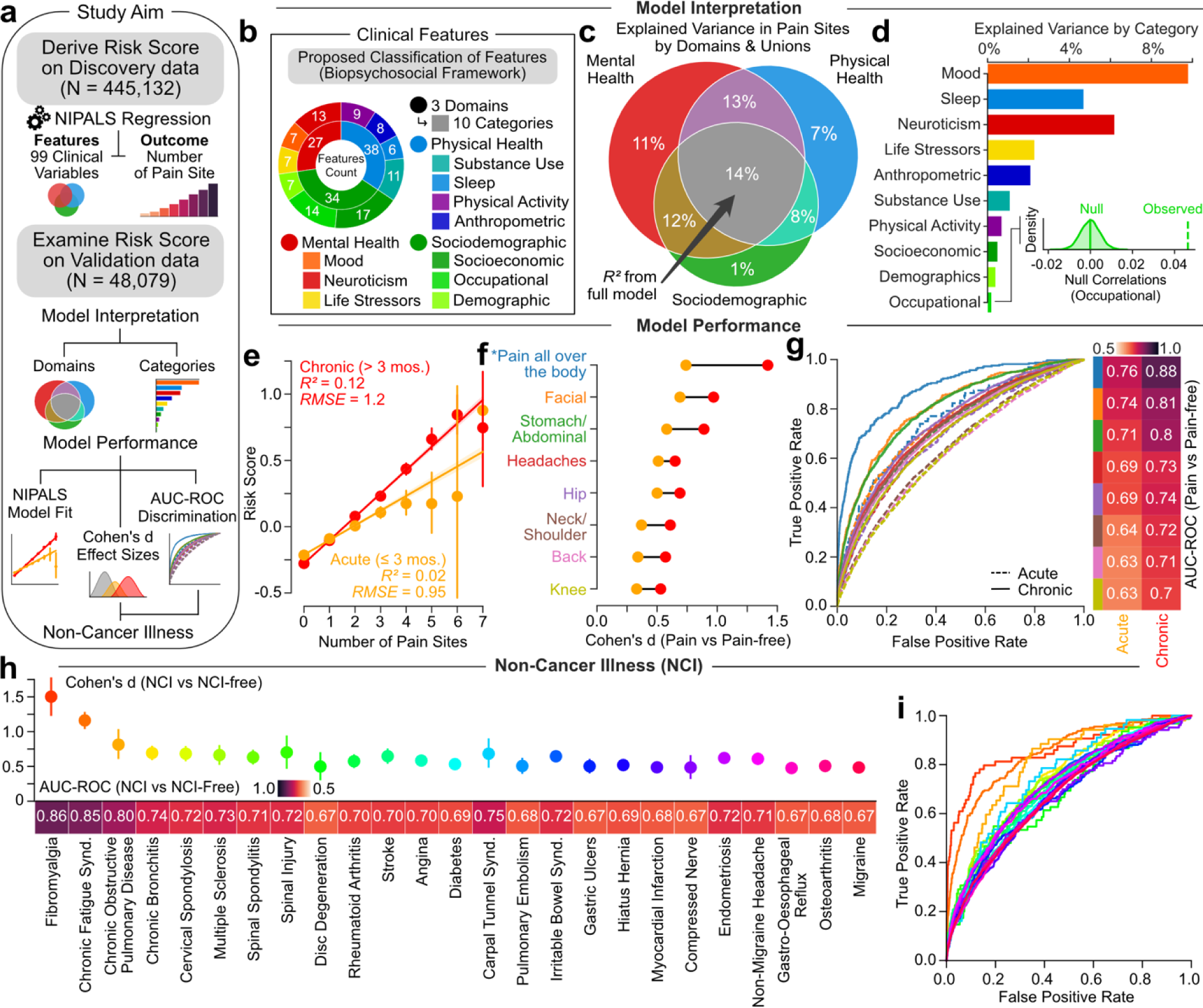
A multivariate model classifying and predicting different pain conditions. **a.** Schematic describing the cross-sectional analyses performed to derive a common risk score predicting the number of pain sites. **b.** Classification of 99 clinical features grouped in 3 domains and 10 categories. **c-d.** Venn diagram and bar graph show the model’s explained variance in the number of pain sites across the 3 domains and the 10 categories. The least contributing category was compared to a null model generated from 10,000 permutations**. e-h.** The model performance is shown in the testing set using explained variance (R^2^) and Root Mean Squared Error (RMSE) for acute and chronic pain conditions separately. Mean estimated across number of sites and standard errors are shown. **f.** Cohen’s *d* effect sizes in the risk score for each pain site (acute in orange and chronic in red) compared to pain- free individuals. **g.** The diagnostic ability of our model to classify acute and chronic pain conditions are displayed using the area under curve of the Receiver Operator Characteristic (AUC-ROC) curve. **h-i.** The diagnostic ability of our model to classify the selected non-cancer illnesses are displayed using Cohen’s d and AUC-ROC (selected NCI compared to NCI-free individuals). Errors bars estimated from 10,000 bootstrap resampling. *Pain all over the body was excluded from model training in the discovery set. Abbreviations: Non-Cancer Illness (NCI); Area Under the Curve (AUC); Receiver Operating Characteristic (ROC).

The strength of the approach is that it considers all the features simultaneously to identifies the optimal solution that best predict the number of pain sites in the cross-validation procedure. The number of co-existing pain sites was mostly explained by mood, sleep, and neuroticism while demographics, and occupational measures explained the least. A detail list of each feature and its respective weight in the model is presented in (**extended fig. 3a**). Notable features with positive weights included tiredness, insomnia, and body mass index, and notable features with negative weights included grip strength, employment status, and frequency of alcohol intake. Although the model was trained irrespective of acute and chronic pain, the number of chronic pain sites was systematically better predicted than the number of acute pain sites (**fig. 2d**). Concordant results were obtained in the discovery set (**extended fig. 3b**). Partial correlations were then used to construct networks at different densities, showing the respective contribution of each of the 10 categories for acute and chronic pain, based on the strength of their conditional associations after controlling for other categories (**extended fig. 3c,d)**. The networks show that chronic pain was simultaneously associated with independent categories, highlighting the multifactorial nature of the model used to predict pain.

**Fig. 3.**
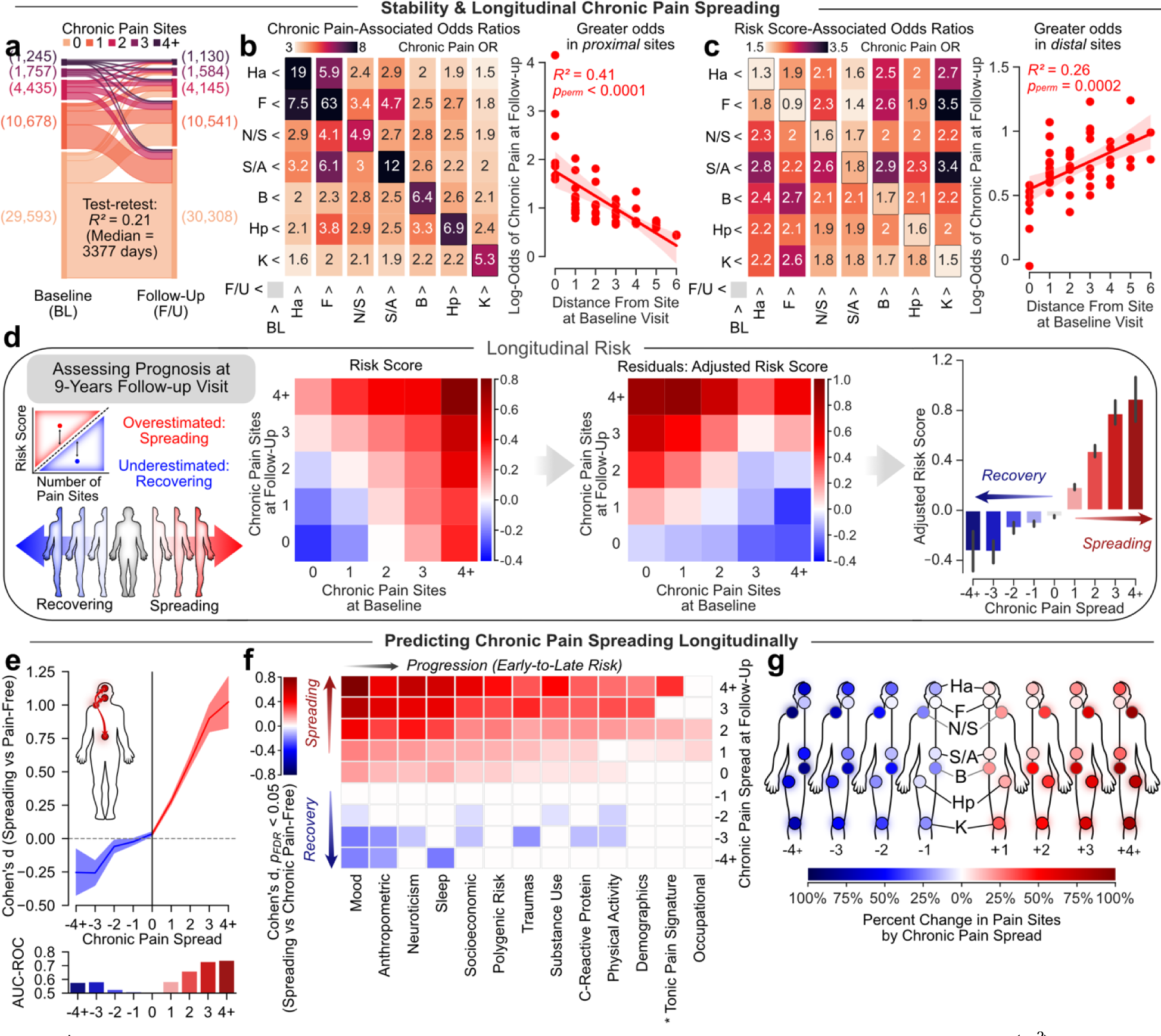
Forecasting the spreading and recovery of chronic pain. **a.** Test-retest variance explained (R^2^) in number of chronic pain sites (4+ including pain all over the body) between baseline and the follow-up visit. **b.** Odds of reporting chronic pain sites at baseline and the follow-up visit depended on the distance on the body map. **c.** our risk score however increased the odds of reporting pain at distal sites. Significance was determined using 10,000 permutations tests. **d.** The matrices display the risk score depending on the changes in the number of chronic pain sites before (left matrix) and after (right matrix) adjusting linearly and exponentially for the number of chronic pain sites initially reported at baseline. A negative adjusted risk score was associated with recovery and a positive adjusted risk score was associated with spreading of chronic pain. **e.** The diagnostic capacities of our adjusted risk score for recovering and spreading was tested using Cohen’s *d* effect size and AUC-ROC discrimination when compared to chronic pain-free participants. **f.** The order of progression between the pain determinants was determined using Cohen’s *d* in each category and biological markers. The factors are ordered depending on their importance in spreading and recovering. Early factors showed significant differences between small changes in chronic pain (Pain +1 or -1) while late factors only showed differences between large changes in chronic pain. **g.** Changes in chronic pain displayed per pain sites. *Tonic Pain Signature was only available at the follow-up visit. Abbreviations: Headache (Ha); Facial (F); Neck or Shoulder (N/S); Stomach or Abdominal (S/A); Back (B); Hip (Hp); Knee (K); Follow-up visit (FU); Baseline visit (BL). Area Under the Curve (AUC); Receiver Operating Characteristic (ROC).

The model’s output provided a single prediction for the number of pain sites for each participant based on their score on the 99 features. This output was referred to as the risk score for pain and was then used to classify each pain condition separately. The risk score for pain showed good to excellent performance for classifying chronic pain conditions from pain free participants at each body site (**fig. 2e,f**; similar performance were observed in the discovery set shown in **extended fig. 4a-d**). The risk score for pain also showed good performance for classifying pain-related clinical conditions diagnosed using the ICD-10 (**fig. 2h,i**). Moreover, the risk score for pain generalized to secondary pain outcomes including overall health rating, use of opioids, and disability due to sickness (**extended fig. 4e-f**). Overall, the risk score for pain generalized well across different chronic pain conditions classified from either their body sites or from the ICD-10 diagnoses.

**Fig. 4.**
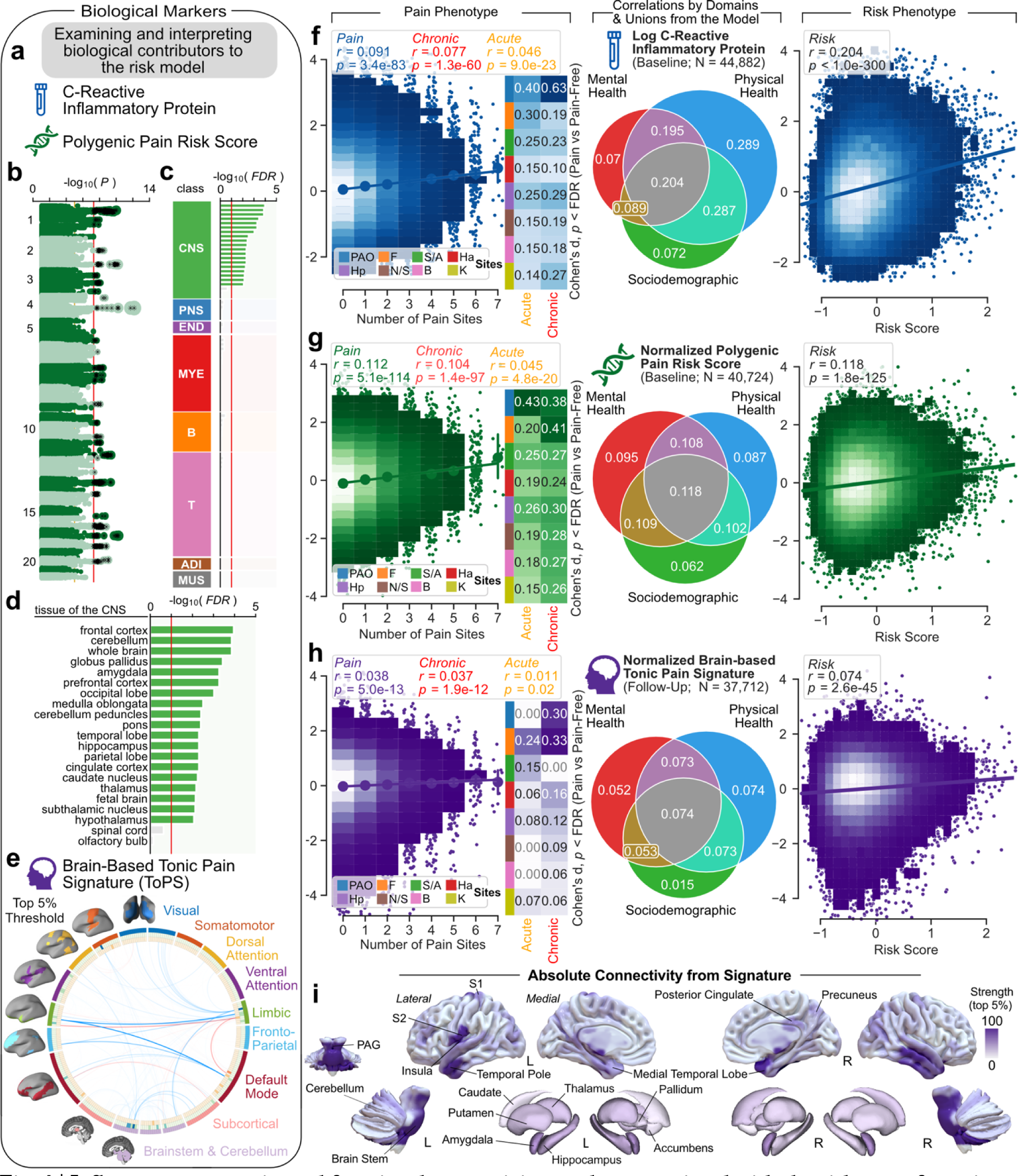
Inflammatory, genetic, and functional connectivity markers associated with the risk score for pain. **a.** Schematic describing the selected biological markers: c-reactive inflammatory protein, a polygenic risk score for the number of pain sites, and a validated brain signature for sustained pain. **b-d.** Genome-wide association study of number of pain sites in the discovery data. **b.** The Manhattan plot shows association p-value for each single nucleotide polymorphism. **c.** The partitioned heritability in tissues of the Benita et al. dataset is shown for total of 78 tissues grouped into eight tissue classes: central nervous system (CNS), peripheral nervous system (PNS), endocrine (END), myeloid (MYE), B cells (B), T cells (T), adipose (ADI) and muscle (MUS). P-values were FDR-adjusted (10%) for enrichment with significant tissues colored. **d.** Details shown for the CNS tissue class**. e.** Circular graph representing the links of the Tonic Pain Signature (ToPS) computed from resting state functional Magnetic Resonance Imaging (rsfMRI) and thresholded for the top 5% of weights. **f-h.** The association between each selected biological marker and the number of pain sites and our risk score. The Venn diagram shows the correlation between the biological markers with respect with the 3 domains. **i.** Rendering of the ToPS brain parcels showing the strongest connections at 5% density. The Cohen’s d effect sizes are presented for each pain site compared to pain-free individuals. Comparisons were FDR-corrected (q = 0.05). Abbreviations: Central Nervous System (CNS); Peripheral Nervous System (PNS); Endocrine (END); Myeloid (MYE); B cells (B); T cells (T); Adipose (ADI); Muscle (MUS); Headache (Ha); Facial (F); Neck or Shoulder (N/S); Stomach or Abdominal (S/A); Back (B); Hip (Hp); Knee (K); Periaqueductal Grey (PAG); Primary Somatosensory Cortex (S1); Secondary Somatosensory Cortex (S2); Left (L); Right (R).

### Predicting recovery and spreading of chronic pain

We used the longitudinal data (*i.e.*, the patients from the test set) to test if the risk score for pain measured at baseline predicted changes in the number of chronic pain sites at the follow-up visit nine years later. The stability and individual changes in the number of pain sites between the two visits are displayed in **fig. 3a**. The matrix in **fig. 3b** shows that chronic pain at baseline was associated with higher odds ratio of experiencing chronic pain at the same site or at a proximal site nine years later. Moreover, individuals with high-risk scores for pain were more likely to report new pain at distal sites (**fig. 3c**). Thus, while baseline chronic pain presents a risk for spreading of pain to proximal sites, higher risk scores for pain instead impacted the spreading of pain to distal sites, where pain is not normally propagating. As hypothesized, the risk score for pain adjusted for the number of pain sites at baseline predispose to the pain outcomes measure at the follow-up visits; patients with negative scores recovered from their pain while patients with positive scores progressed towards a spreading of their pain (**fig. 3d**). Therefore, our adjusted risk score shows strong effect sizes and obtained good performance for predicting chronic pain spreading over multiple new pain sites at the follow-up visit (**fig. 3e**).

We next performed a tentative temporal ordering of individual risk factors by ranking the 10 categories and the three pre-selected biological markers for pain based on their effect sizes. This allowed us to unpack the sequence of biopsychosocial risk factors, from early prodromal features to late features, predicting the progression of the spreading or recovering of chronic pain across body sites (**fig. 5f**). Here, mood was the earliest contributor to the spreading of pain, biological contributors were middle to late onset risk factors, and occupational ranked last. The changes in chronic pain used in the temporal ordering analysis are displayed per body sites in **fig. 5g**.

**Fig. 5.**
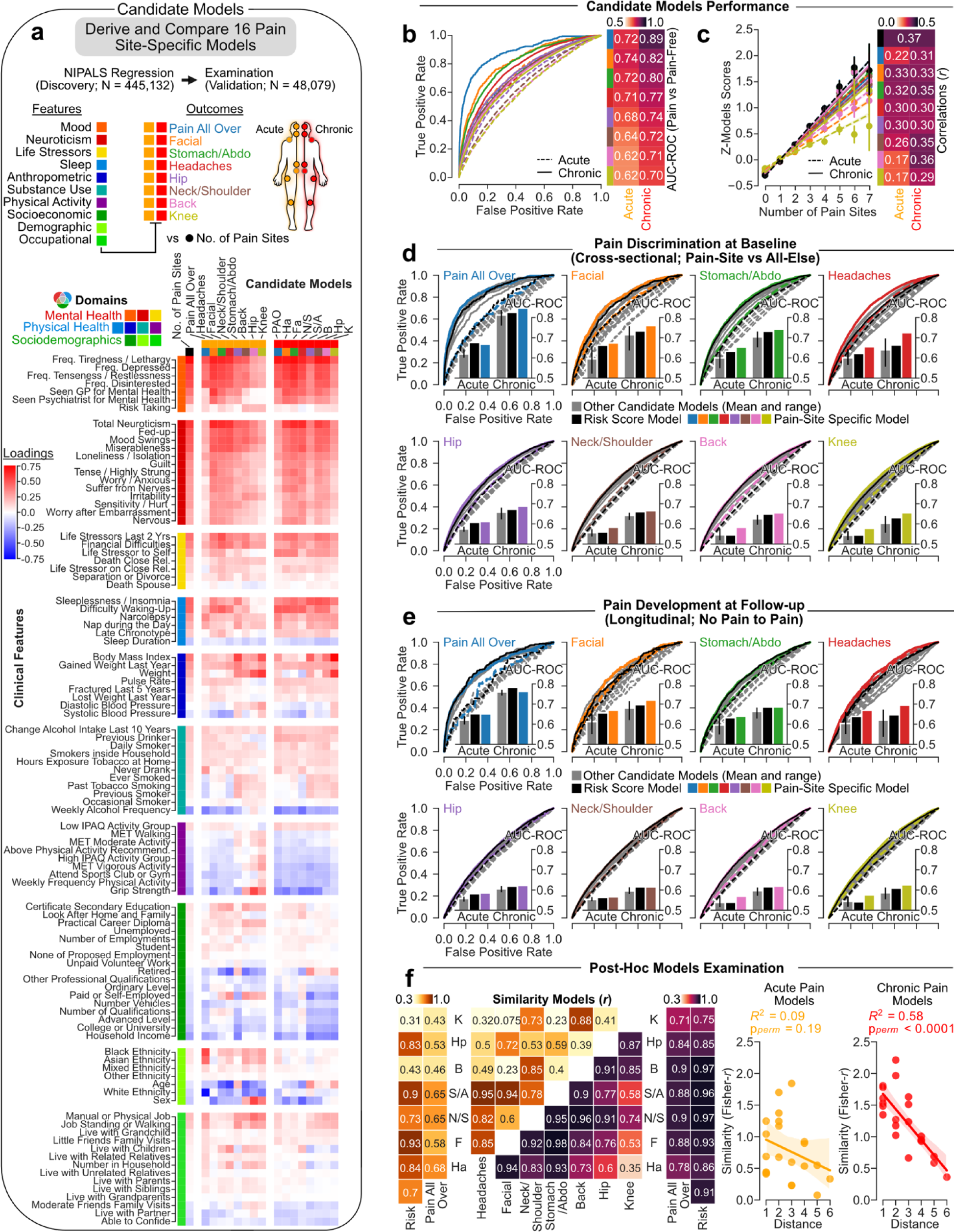
A common risk shared across chronic pain conditions. **a.** A schematic describing that a total of 16 site- specific candidate models (e.g., acute knee vs all-else) were derived cross-sectionally in the discovery set using NIPALS. Features loading (Pearson’s r correlation coefficient between features and the models’ scores) are shown in the testing set for each model. **b.** Candidate models’ capacities to discriminate between the pain sites they were trained on from pain-free individuals are shown using the Area Under the Receiver Operator Characteristic Curve (AUC-ROC). **c.** The risk score derived from each candidate model correlated with number of co-existing pain sites. **d.** Cross-sectional discrimination for each pain site in acute (dashed line) and chronic (full line) pain conditions against the rest of the testing cohort (i.e., pain-free and other pain sites) using the model specific to the site (in color), to the number of pain sites (black), and to other candidate models trained on different pain sites (grey). **e.** The same analyses were performed in the longitudinal data to predict the development of chronic pain in pain free individuals 9-years later. **f.** Post-hoc analyses show that similarities between the feature loadings of the different models are associated with the distance between the sites for chronic pain conditions. Abbreviations: Headache (Ha); Facial (F); Neck or Shoulder (N/S); Stomach or Abdominal (S/A); Back (B); Hip (Hp); Knee (K); Area Under the Curve (AUC); Receiver Operating Characteristic (ROC).

### The risk score for pain is embedded with biological markers

The association between the risk score for pain and biological markers of pain was further examined in the participants of the testing set (**fig. 4a,b**). The selected biological markers included an inflammatory blood marker (c-reactive protein; CRP), a Polygenic Risk Score (PRS), and a validated brain-based biomarker for pain (Tonic Pain Signature; ToPS ^25^). Here, the PRS was computed from a GWAS analysis for the number of pain sites in discovery set **fig. 4c**. We found significant enrichment for heritability at loci of genes expressed most specifically in brain regions. All biological markers were significantly associated with the number of pain sites (**Fig. 4f-h**), although the magnitude of the correlations was smaller than expected, especially for the brain-based signature. Yet, the inflammatory blood marker and the brain-based biomarker were more strongly correlated with our risk score for pain than with the actual number of pain sites, emphasizing that psychosocial factors were embedded with the biological factors for pain. Findings from the discovery set and additional exploratory analyses for the biological markers are shown in **extended fig. 5-7**.

### Common risk scores for different chronic pain conditions

We next investigated the specificity of the risk factors between different pain conditions by generating and testing alternative candidate models for each pain site separately (**fig. 5a**). Sixteen new candidate models were trained by applying the NIPALS algorithm on the 99 features to classify each body site separately (e.g., patients reporting chronic knee pain versus everyone else). Here again, the results were properly validated in the left-out participants of the testing set. The matrix presented in **fig. 5a** shows the loadings of the 99 features (rows) on the risk score derived for different pain conditions (columns), including our initial model predicting the number of co-existing pain sites (black). A visual inspection of the matrix show that the most predictive features were also the most homogenous, from mental health, physical health and sociodemographics. For example, different loadings were observed for some demographic features such as age, sex, and ethnicity, some socioeconomic features such as retiring, and self-employment, and the grip strength feature depending on the pain conditions. The models trained to classify acute pain conditions showed poor to good discrimination (AUC ranging between 0.62-0.74) while the models trained to classify chronic pain conditions showed good to excellent performance (AUC: 0.70-0.89; **fig. 5b**). The expression of each risk score (normalized for comparisons) correlated with the number of co-existing pain sites (**fig. 5c**). The weights of the 99 features are presented for each candidate model trained for different pain conditions in **extended fig. 8a,b**.

To test the commonality or the specificity between the body sites, we compared the discriminative value of each site-specific model with that of the candidate models trained on a different pain site, in both cross-sectional (**fig. 5d**) and longitudinal (**fig. 5e**) data. In the cross-sectional data, site-specific models showed only modest improvement over the candidate models trained for a different body site. In the longitudinal data, we applied these models to predict development of chronic pain conditions in individuals that were pain free at baseline. Here again, site-specific models showed only modest improvement over candidate models trained on another pain site. Similar results were observed in the discovery dataset (**extended fig. 8c,d**). We conclude that different chronic pain condition can be predicted from interchangeable models trained on a different pain site. This finding supports the proposition that a single biopsychosocial framework may be used to characterize a general *chronic pain syndrome*.

To further examine this finding, we generated a similarity matrix correlating the loadings between the candidate models. The weights of our initial model trained on the number of pain sites strongly correlated with the weights of the eight candidate models trained for chronic pain conditions (*r* = 0.68-0.94; **fig. 5f**). Moreover, the similarities between the chronic pain candidate models depended on the distance between the body site, reflecting the actual body map of the co- existing pain sites presented in **fig. 1b**. This was however not the case for acute pain conditions, as the candidate models were dissimilar from one another, less discriminable, and did not depend on the distance on the body map. Similar results were obtained in the discovery dataset (**extended fig. 8c,d**).

### A sparse model for chronic pain

Lastly, we aimed at simplifying our model by reducing the number of features, as a sparse model would be more useful in research or clinical settings. A six items model using sleep neuroticism, mood, trauma, and anthropometric measurements (**fig. 6a,b**) achieved good performance in cross-sectional data, average to good performance in longitudinal data (**fig. 6c,d**), and predicted the impact of pain for the patients (**fig. 6e**). This represented the best trade-off between the smallest number of features and the highest AUC-ROC, especially in the longitudinal data. This risk score for pain was calculated by simply summing the binarized answers to the 6 items. None of these features were directly related with pain or attitude towards pain, suggesting that more objective predictions can be obtained by avoiding the use of pain questionnaires to predict pain outcomes.

**Fig. 6.**
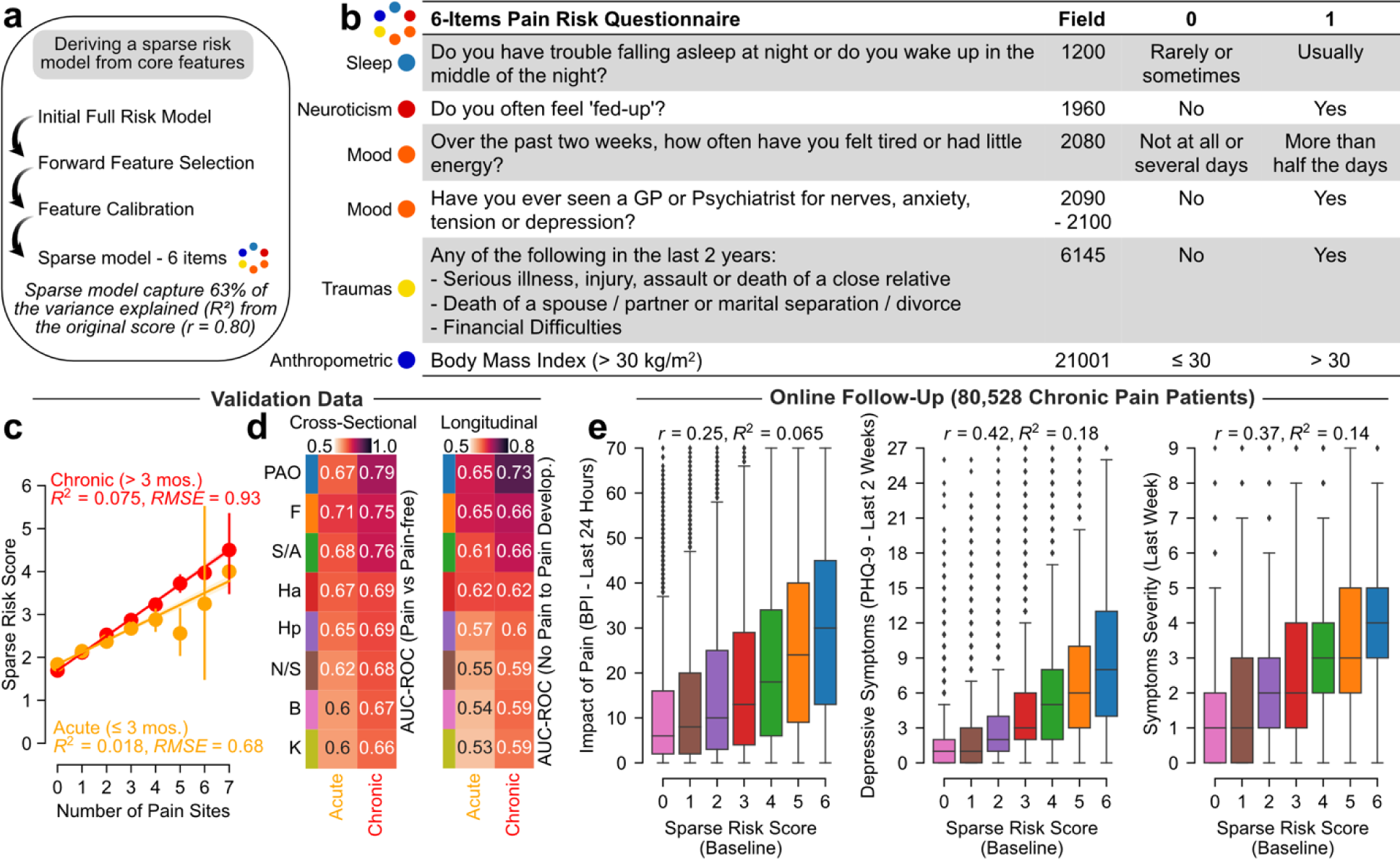
Sparse model for chronic pain. **a.** Schematic describing the steps implemented to derive a sparse model. **b.** Core selected feature retained and binarized to form a 6-items short questionnaire capturing 63% of the variance explained by the full risk score predicting the number of pain sites. **c-d.** Model performance on the testing set for both acute and chronic conditions and in cross-sectional and longitudinal data. **e.** In the online follow-up pain questionnaires, the sparse model was associated with the interference of pain, the depressive symptoms severity in last 2 weeks, and symptoms severity during the last week. Abbreviations: Headache (Ha); Facial (F); Neck or Shoulder (N/S); Stomach or Abdominal (S/A); Back (B); Hip (Hp); Knee (K); Brief Pain Inventory (BPI); Patient Health Questionnaire (PHQ).

## Discussion

A major challenge in understanding chronic pain conditions is that they are heterogeneous by nature and commonly overlap ^19, 26^. For the first time, we provide empirical evidence for a biopsychosocial risk score that can predict pain severity and differentiate patients experiencing chronic pain from healthy controls. Here, we studied pain conditions through the lens of a continuum, where pain was examined in either the acute or chronic phase, from the body site where it is experienced, from the medical diagnoses, and from the number of co- existing pain sites. Our model, trained to predict the number of pain sites, could classify chronic pain conditions at different body sites, general health, usage of opioid, disability, and 25 Non-Cancer Illnesses. Importantly, we show that the risk score could forecast the spreading or recovery of chronic pain nine years later, in a group of new patients. The ability to forecast chronic pain conditions can be used in many contexts, including selecting patients in research protocols, matching individuals between the arms in randomized controlled trials, or guiding more aggressive treatment options in patients requiring urgent clinical attention.

For over 40 years, the biopsychosocial model has emphasized that illnesses are determined from a synergic interaction between psychological, sociocultural, and biological factors ^27^. The biopsychosocial model has been influential in the field of chronic pain as any model focusing solely on any one of these domains would inevitably be incomplete or inadequate ^5^. Here, we investigated how psychosocial risk relates with biological markers of pain. The three pre-selected biological factors (i.e., CRP, PRS, and ToPS) showed small but consistent associations with the number of pain sites (r = 0.04-0.11). Previous studies have demonstrated that brain wide associations ^14^ or genome wide associations ^15^ performed in well powered studies are smaller than those initially observed in small sample studies. In the present study, the CRP, PRS, and ToPS were tested in more than 30,000 individuals, making it safe to conclude that their associations with pain were small. Interestingly, these biological factors more strongly correlated with the risk score than with the number of pain sites *per se*. Tentative temporal ordering also revealed earlier elevated psychosocial risk (mood, anthropometric, neuroticism) compared to biological risk developing significantly in patients with higher numbers of pain sites. These findings suggest that biological factors may be more strongly expressed in the risk score predisposing chronic pain development, raising questions regarding the pathophysiological nature of chronic pain illnesses in these patients. This raises questions regarding the utility of the examined biological markers for risk given the sensitivity of psychosocial risk to detect pain in lower numbers of pain sites.

Although the origin of pain vastly varies between pain conditions and taxonomies, we further demonstrated that chronic pain is determined from common risk factors. Body-site specific models for chronic pain risk showed little superiority over site agnostic and even off-site models. Although unexpected, this finding again suggests that different chronic pain conditions can be predicted cross-sectionally and longitudinally from a common psychosocial framework. Mood (*i.e.,* consulting GP for depression or anxiety), sleep (*i.e.,* insomnia), neuroticism (*i.e.,* feeling fed-up), and anthropometric measures (BMI) were the major risk factors identified in this study. Interestingly, our risk score and candidate models showed that acute pain conditions were harder to predict than chronic pain conditions. The strongest predicting features to number of co- existing pain sites, mental health related features, showed homogeneity across sites while the least predicting features, sociodemographics, showed heterogeneity. Overall, we found little evidence of pain-condition specific psychosocial risk factors. Instead, we further demonstrate that risk factors support a general pain syndrome with a common biopsychosocial framework. While it has been known that the biopsychosocial framework can predict the experience and maintenance of pain, we found here that this aggregated risk could predispose the development of new pain sites, a phenomenon we refer to as the “spreading” of pain sites.

An increasing number of conditions and syndromes resembling widespread pain disorders have been referred as chronic overlapping pain conditions (COPC). We explore these co-morbidities by looking at multiple co-occurring pain sites beyond the sites of focus (beyond headaches pain in migraine, stomach/abdominal for IBS, or hand pain for carpel tunnel syndrome) to demonstrate this co-occurrence of pain sites to be common in other conditions than the traditional COPC. Furthermore, we demonstrated that the co-occurrence of pain sites was not random, with a strong independence between proximal pain sites, shown from acute and chronic pain sites and correlations between pain intensity ratings. More interestingly, pain between distal sites, high spreading of pain, and fibromyalgia patients were best predicted by biopsychosocial risks, suggesting that elevated risk could present a pathway to the progression of widespread pain disorders and may help us understand how one site can spread to multiple.

Our model also identified the features of a common driver that can be referred to as a general *chronic pain syndrome* that would encompass what has previously been referred to as *high impact pain*. High impact pain has previously been used to define patients who are living with limitations in work, social, and self-care activities leading to disability, opioid use, and needs for healthcare resources ^28, 29^. While our framework was derived to predict the number of co-existing pain sites, it could also predict the psychological and social limitations considered key indicators of pain impact. This denotes that the concepts of chronic pain spreading and high impact pain appear intimately linked and predictable from higher order characteristics presented from our biopsychosocial framework.

In conclusion, our model predicted chronic pain spreading across multiple body sites in 50,000 out-of-sample individuals that attended both the baseline visit and the brain imaging follow-up visit nine years later. Data reduction strategies showed that high sensitivity and specificity can still be obtained for some chronic pain conditions after reducing the number of features to six questions. The proposition that chronic pain is a general syndrome that can be accurately predicted with minimal effort has the potential to improve research protocols, clinical practices, and randomization of participants in clinical trials.

## Data Availability

All data are provided from the UK Biobank and available to other investigators online upon permission granted by www.ukbiobank.ac.uk. Restrictions apply to the availability of these data, which were used under license for the current study (Project ID: 20802).

## Methods

### UK Biobank population

The UK Biobank is a prospective epidemiological cohort of approximately 500,000 middle-aged and older individuals across Great Britain (40-69 years old at recruitment). Participants provided electronically signed consent, answered socio- demographic, lifestyle, and health-related questionnaires, and completed a range of physical measures ^1^. A subset of approximately 50,000 participants underwent an additional follow-up visit 6-10 years later (median = 9 years) which included whole-body MRI imaging. The present analyses were conducted under the UK Biobank application 20802. All participants provided written, informed consent, and the study was approved by the Research Ethics Committee (REC number 11/NW/0382). Further information on the consent procedure can be found elsewhere (http://biobank.ctsu.ox.ac.uk/crystal/field.cgi?id=200).

### Pain status at baseline visit

Participants were asked if they experienced pain in the past 3 months interfering with their usual activity at any of the following major anatomical sites – head, face, neck or shoulder, back, stomach or abdominal, hip, knee, or pain all over the body (data field 6159). Note that if *pain all over the body* was selected, no other pain sites could be selected. When answering positively to a given site, participants were asked if the pain lasted more than 3 months. This question was used to distinguish between a chronic pain site, one present for more than 3 months according to the classification from the International Association for the Study of Pain (IASP ^2^) and an acute pain site, one present for less than 3 months. Finally, the number of the pain sites experienced (excluding pain all over the body) was used to measure the co-existing pain sites (i.e. widespread pain) as a continuum. The body sites included in the UK Biobank remained the most prevalent and stereotypical sites of chronic pain.

### Co-occurrence

The co-occurrence of acute and chronic pain sites was measured using odd ratios from the exponential function of a logistic regression coefficient estimated for each combination of sites (excluding *pain all over the body*). The number of sites setting apart each combination was used a measure of their distance (1-7), as done by Khoury et al. (2022) ^3^. To ensure the robustness of the association between co-occurrence and distance, our results were compared to a null model generated from 10,000 permutations tests, using two-sided tests.

### Non-cancer illnesses

The prevalence of pain sites in the UK Biobank among self-reported non-cancer illnesses was also assessed. Patients were asked during a verbal interview if they had been told by a doctor about any serious illnesses or disabilities. If the participant was uncertain of the type of illness they had, they would describe it to the interviewer (a trained nurse) who would attempt to place it within the coding tree (data field 20002).

### Online Follow-up

Online questionnaires were administered about 8-13 years after the baseline visit to better phenotype chronic pain patients. A total of 167,000 participants filled out the *experience of pain questionnaire* (10 sections; https://biobank.ctsu.ox.ac.uk/crystal/crystal/docs/pain_questionnaire.pdf) administered online by the UK Biobank. About 90,000 of these individuals reported pain or discomfort that has lasted for more than 3 months (i.e. chronic). In this online questionnaire, various subsections were used to validate the importance of increased widespread pain using the same major anatomical sites assessed in person. These included the duration of the pain or discomfort (3-12 months, 1-5 years or more than 5 years), worst pain rating experience in the past 24 hours, pain interference using the Brief Pain Inventory across 7 dimensions – each out of 10 in the last 24 hours (10 = as bad as you can imagine; BPI ^4^), depressive symptoms in the last 2 weeks using the Patient Health Questionnaire (ratings of 5-9 ratings associated with mild depression; PHQ-9 ^5^), and severity of pain symptoms in the past week combining three rating scales on fatigue, sleep quality and cognitive symptoms (3 = pervasive, continuous, life disturbing problems). Online follow-up also included examination of 14 chronic pain diagnoses as well as questions about pain including additional anatomical sites and ratings at each site. This allowed us to validate the co-occurrence of pain sites and explore the interdependence of pain ratings accompanying the co-occurrence of pain sites.

### Risk score for pain

A total of 99 features collected at baseline were selected a-priori based on their relevance to chronic pain. The selection was based on the Prognosis Research Strategy (PROGRESS) group who recently provided a framework for the development of a prognostic model to determine risk profile ^6^. Variables were organized along a hierarchical framework from 99 variables into 10 categories forming three distinct domains (mental health, physical health, and sociodemographic). The mental health domain included 3 categories: i) neuroticism – all individuals items and their sum-score – based on neurotic behaviors closely linked to negative affect, ii) stressful life events – illness, injury, bereavement, or stress in the last 2 years – including 6 events, and iii) mood – reported frequency of certain moods in the past 2 weeks and visits to a general practitioner or psychiatrist for nerves, anxiety, tension, or depression. The physical health domain included 4 categories: i) physical activity – Metabolic Equivalent Task scores computed using the International Physical Activity Questionnaire guidelines ^7^, ii) sleep – questions regarding duration, napping, snoring, sleeplessness, iii) substance use – smoking and alcohol use, and iv) anthropometric measures – body mass index, fractures that occurred over the last 5 years, blood pressures, etc.. The sociodemographic domain included 3 categories: i) socioeconomic status – education completed, income, employment, etc., ii) occupational measures – individuals present within household, social entourage, and manual or physical job, and iii) demographic – age, sex, and ethnicity.

### Data exclusion

Among the 99 features selected, participants with more than 20% of features missing were excluded, as were participants with missing data at any of the acute or chronic pain sites (<2.5% of the population). To ensure the findings of the study to be as generalizable as possible to the greater population, no other exclusion criteria were applied. This led to the discovery cohort of 445,132 participants who did not attend a follow-up visit and from whom the multivariate model was derived. The validation cohort of participants who did attend a follow-up visit included 48,079 participants. To minimize potential bias from incomplete questionnaires, a data-driven Bayesian ridge regression model was applied for imputation of missing data as a function of all other features in the model, using the median as prior. A median-only feature imputation was also tried and presented congruent results. Features were then standardized across the participants by centering mean to zero and scaling the variance to one. The same process of exclusion followed by imputation for missing data and standardization with the same mean and variance was applied separately for the validation dataset.

### NIPALS

Finally, a Nonlinear Iterative Partial Least Square (NIPALS) regression algorithm (implemented using scikit-learn.org/) was used to derive an epidemiological model that explained the number of pain sites reported at the baseline visit. This model excluded individuals reporting *pain all over the body* to avoid making assumptions about the equivalence of *pain all over the body* and some number of pain sites experienced. This specific algorithm was chosen to reduce the 99 features into a few sets of distinct homogenous components associated with self-reported number of pain sites. A common rule-of-thumb in multiple regression suggests that the minimum ratio of sample size per variable is 10:1 with greater ratios equivalent to greater stability. Here, we observe a 4500:1 ratio of sample size per variable giving us confidence in our stability. Ten-fold cross-validation was used to assess the number of components to use in the model. A total of three components were selected based on the largest increase in the variance explained and the largest decrease in root mean squared error according to the elbow criterion. Then, the model was then applied in the validation dataset.

### Evaluation of the model

The contribution of the domains and categories were examined using the weighted sum of the features across each category, domain or the union between domains. Model fit to number of pain sites was assessed using explained variance and root mean squared error. To examine the impact of acute and chronic pain sites, the model score of each pain site was compared to the score of pain-free participants. Group differences were computed using both Cohen’s d for effect size (pooled standard deviation) and the Area Under the Receiver Operating Characteristic curve (AUC-ROC) for discrimination. AUC-ROCs were used to estimate model accuracy because they are i) threshold-unspecific and ii) resilient to class imbalance, which is inherent to less frequent pain conditions or clinical outcomes. The performance of the model was also tested across 25 different Non-Cancer Illnesses (NCIs) commonly associated with chronic pain using the same metrics: Cohen’s d and AUC-ROC. To ensure the robustness of the results in less frequent NCIs, 10,000 bootstraps resampling were done to estimate the confidence interval in the observed effect sizes.

### Network analyses

Networks were constructed from partial correlations between pain and the 10 categories. Partial correlations were used to measure conditional dependence between categories – defined as nodes – while controlling for all other potential edges. The number of acute pain sites and chronic pain sites were integrated into the network to assess the relative contributions of our model’s categories on both pain types. The networks were constructed and studied at three different densities. A threshold was first applied to obtain a sparse model and conserve connections equivalent to a small effect size (partial correlation > 0.1). An intermediate model was then constructed using a more liberal threshold equivalent to a very small effect size (partial correlation > 0.05). A full model was finally constructed by including all the edges surviving Bonferroni-correction. Nodes were placed using a force-displacement layout (i.e. spring layout, using Fruchterman-Reingold algorithm) using *qgraph* ^8^. Starting in a circular layout and through various iterations, more connected nodes are placed closer together while less connected or negatively linked nodes are placed further apart. Finally, node-weighted centrality, a measure of the mean number of edges passing through each node, was computed to estimate the centrality of both categories and pain outcomes. The procedure was done on both the discovery and validation datasets.

### Secondary pain outcomes

We examined overall health rating (field 2178), opioid medication use (NO2A from the ATC classification), and inability to work due to sickness or disability (field 6142). Opioid medication use was obtained from self-reported of medications at each visit (data field 20003), and was coded according to the Anatomical Therapeutical Chemical (ATC) classification system from the World Health Organization obtained from the Wu et al. (2019) classification ^9^. The performance of the model in predicting longitudinal changes in opioid use and changes in ability to work were also tested.

### Chronic pain spreading

The risk score for pain was initially derived using cross-sectional data. The prognostic value of the pain risk score to predict the development, persistence, and worsening of chronic pain was however assessed using the left-out participants in the validation for who the longitudinal data were available. After examining the stability of number of pain sites (0 – 4 or more including pain all over the body), we measured the risk of chronic pain at each anatomical site at follow-up associated with chronic pain at each site at baseline. Odds ratios using the exponential function of the logistic regression coefficients were used to calculate risk. The risk of chronic pain at each anatomical site at follow-up was also examined by calculating the odds ratio associated with one unit higher in the risk score for each chronic pain site at baseline.

Spreading was measured using the change in number of chronic pain sites (from -4 or less to 4 or more). To examine the prognostic value of our risk score, we regressed out the number of chronic pain sites (and their squared values) from the risk score at baseline making the score orthogonal to the pain at baseline. Making the score orthogonal allowed us to interpret interindividual deviations in this adjusted score as risk of recovery or spreading of pain. Group differences in the adjusted risk between individuals without chronic pain and individuals with chronic pain presenting different rates of pain spreading or recovery were computed using AUC-ROC discrimination and Cohen’s d effect sizes.

A temporal ordering of predicted risk across the derived ten categories and three biological markers adjusted for the number of pain sites (and squared values) was performed at baseline. Difference in predicted risk across each rate of chronic pain spread compared to chronic pain-free individuals for all 13 factors were computed using Cohen’s *d* effect size while significance was corrected for false discovery rate across all rates of spreading and factors. Events were ranked according to the absolute sum of effect sizes across rates of spreading providing a temporal progression of the spreading or recovery of chronic pain across body sites. The associated changes for each rate of spreading were also measured using the sum of chronic pain sites developed (+1) or recovered (-1) across the rate of chronic pain spread or recovery to investigate early-to-late pain site development.

### Immune-inflammatory profile

The UK Biobank haematological data included a complete blood count (https://biobank.ctsu.ox.ac.uk/crystal/crystal/docs/haematology.pdf). The sample handling and storage has been described by Elitt & Peakman (2008) ^10^. Inflammation was estimated using C-Reactive Protein (CRP; field 30710) obtained through saliva samples and measured by immunoturbidimetric assay using a high sensitivity analysis on a Beckman Coulter Analyzer. A logarithmic transformation was applied to the raw measures of CRP to account for the positive skewness (https://biobank.ndph.ox.ac.uk/showcase/field.cgi?id=30710). Immune cell count included neutrophils, platelets, reticulocytes, basophils, lymphocytes, eosinophiles, and monocytes, most of which have been shown to be independently linked to chronic pain, the sickness response and associated depressive profile ^11^.

### Polygenic Risk Score

A genome-wide association study (GWAS) of number of pain sites, including both acute and chronic was conducted. A thresholding procedure was done across 7 statistical thresholds of significance (from P = 5x10^-2^ to 5x10^-8^) for each single nucleotide polymorphism. The association of each threshold with the risk score, c-reactive protein and pain phenotype was also examined in the discovery and validation datasets.

Partitioned heritability in tissues was used to investigate the genetic architecture of our polygenic risk score. The top 1,000 most enriched genes per tissue were extracted from the gene expression databased features in Benita et al. (2010) using the computer program ‘ldsc’ ^12–14^. A total of 78 tissues grouped into eight tissue classes – central nervous system, peripheral nervous system, endocrine, myeloid, B cells, T cells, adipose, and muscle – were examined for enrichment. The methodology is described in greater detail in our previous publication ^3^

### Tonic Pain Signature (ToPS)

The resting state functional Magnetic Resonance Imaging data were requested to the UK Biobank to construct the brain network from which the ToPS was derived. Our study used the minimally preprocessed pipeline designed and carried out by FMRIB group, Oxford University, UK ^15^. The minimally preprocessed resting-state fMRI data from the UK Biobank were analyzed using the following preprocessing steps: motion correction with MCFLIRT ^16^, grand-mean intensity normalization, high pass temporal filter, fieldmap unwarping, and gradient distortion correction. Noise terms were identified and removed using FSL ICA-FIX. Full information on the UK Biobank preprocessing is published ^15^. Additional preprocessing included warping the image in native space to the 2mm MNI template (FSL), despiking using 3DDespike (AFNI from Nipype), 6-mm kernel smoothing (Nilearn), and resampling to 3-mm (for storage purposes). A modified Brainnetome atlas ^17^ was used to parcel the brain into 279 distinct regions to apply the weights from the Tonic Pain Signature (ToPS ^18^), a capsaicin-induced tonic pain signature of pain derived from the brain that was associated with both experimental and clinical pain. The modified atlas includes additional midbrain, brainstem, and cerebellar regions.

Dynamic connectivity was estimated to derive ToPS using Dynamic Conditional Correlation (DCC). DCC is based on generalized autoregressive condition heteroscedastic (GARCH) and exponential weighted moving average (EWMA) models (implemented by https://cocoanlab.github.io/tops/). The preprocessing aimed to be as similar as possible to the original ToPS study without diverging from the minimally preprocessed data from the UK Biobank. The weights of the signature were thresholded to the top 5% to avoid overfitting and to minimize relation with head motion prior to the examination of the full dataset (early subsample of n = 200). Multiple thresholds (1, 2.5, 5, 10, 25, 50, 100%) were also tested to ensure generalizability. Absolute connectivity from the signature for visual and interpretation purposes was computed using the sum of absolute connectivity values for each brain region within the top 5% threshold (z-scores normalized prior to thresholding) using a cut-off of 100 as the maximum. Surface rendering was done using the *Surf Ice* tool (https://www.nitrc.org/projects/surfice/).

Two frameworks were evaluated to control for the effects of confounding variables: i) adjusting confounding variance that overlaps with pain and ii) adjusting total confounding variance. The first approach allows the brain signature to be compared to other polygenic and inflammatory markers that were left intact given the research focus on prediction, while the second ensures that our results did not overlap with confounds commonly reported as higher in pain patients such as motion. Results were very similar in both approaches with the former presenting slightly smaller probability values.

The MRI-based covariates included head motion (linear, squared, and cubed), imaging site, position in the scanner, and coil position (X, Y, Z respectively). Both covariates and brain features were normalized to a mean of zero and variance of one unit across participants. To examine confounding variance that did not overlap with pain, the number of pain sites was regressed out from confounds. A confound-removal procedure, done on the original confounds or pain-regressed confounds was applied by deriving a multivariate regression model to predict each normalized brain feature as a function of the normalized confounds. The procedure was done for each of the brain features, making them strongly or perfectly orthogonal to confounds.

### Candidates Models

The same statistical procedure (NIPALS) performed on the same the 99 clinical features was used to derive our initial model predicting the number of pain site and the 16 candidate models classifying acute or chronic pain sites (vs all-else). Model specification was done through 10-fold cross-validation to maximize the variance explained (R^2^) while minimizing the Root Mean Squared Error (RMSE). This allowed us to decide on the sparsity of components to include in the models using the elbow criterion from the largest drop of explained variance. The same parameters (i.e. three components, used as regularization) were used to predict the pain sites using NIPALS. Features for each model were visualized using two methods: i) by computing the Pearson’s r correlation equivalent to the loadings of each feature onto their projected score or ii) by comparing the z-normalized weights used to obtain the projected score. The former approach was preferred for interpretability.

The sensitivity of these candidate models was evaluated using AUC-ROC discrimination in comparison to pain-free individuals and Pearson’s R correlation with number of pain sites. Then, we assessed the specificity of these models by comparing their AUC-ROCs to the AUC-ROC of i) the risk score model and ii) the pain-site models of same origin (e.g. acute knee pain). This was done cross-sectionally (e.g. acute knee pain vs all-else) and longitudinally for the risk of developing a given pain sites in individuals without it.

Finally, a post-hoc analysis was done to examine the similarity across model using Pearson’s r correlation coefficient between the 99 loadings (or normalized weights) obtained in acute and chronic models. This approach allowed us to compare the similarity between risk factors estimated from each model. These correlations were then normalized using z-fisher transformations to estimate the association between similarity of pain sites models and the distance between pain sites.

### Sparse model for pain risk score

A sparse model containing 6 features was derived from the full risk model containing 99 features. We trained a linear forward feature selection algorithm to select the core six features that presented the highest explained variance based on the full risk model. Forward feature selection iteratively evaluates a prespecified combination of six features in a feature pool (99 from the original model) until there is no improvement in the model’s performance. The feature selection algorithm chose features spanning across 5 categories (Sleep, Neuroticism, Mood, Life stressors, and Anthropometric).

Core features were binarized and calibrated based on feature thresholds that maximized the discrimination performance (AUC-ROC) between subjects reporting pain all over the body in the online follow-up data and those not reporting pain all over the body (n = 159,663). This led to the formation of a 6-item short questionnaire capturing 63% of the variance explained by the full risk model. The cross-sectional and longitudinal performance of the sparse model was assessed in the discovery set for acute and chronic conditions using the number of pain sites (explained variance and root mean squared error) and group differences between pain and pain-free groups (AUC-ROC). The sparse model was also evaluated on the online follow-up pain questionnaires in a subsample of 80,000 participants on interference of pain ratings (BPI, depressive symptoms in the last 2 weeks, and the severity of symptoms using Pearson’s correlation and R-squared metrics.

### Data Availability

All data are provided from the UK Biobank and available to other investigators online upon permission granted by www.ukbiobank.ac.uk. Restrictions apply to the availability of these data, which were used under license for the current study Project ID: 20802).

### Code Availability

Detailed code and annotation will be published upon publication and can be found on GitHub (LINK). The medication classification done by Wu et al. (2019) can be found in supplementary from the original article (https://www.nature.com/articles/s41467-019-09572-5). Code to extract the ToPS by Lee et al. (2021) can be found online (https://cocoanlab.github.io/tops/)

## Acknowledgement

This work was supported by the Canadian Institutes of Health Research (CIHR 453096), the Louise and Alan Edwards Grants in Pain Research. C.T.S. was supported by the Healthy Brains Healthy Lives (HBHL) initiative (Canada First Research Excellence Fund) and by the Elaine Bélanger Graduate Fellowship in Medical Research (Faculty of Medicine and Health Sciences). M.F. was also supported by the HBHL initiative. This study makes use of data from UK Biobank (Project ID: 20802) and we thank the UK Biobank participants and the UK Biobank team for generating an important research resource.

---

**Extended Fig. 1.**
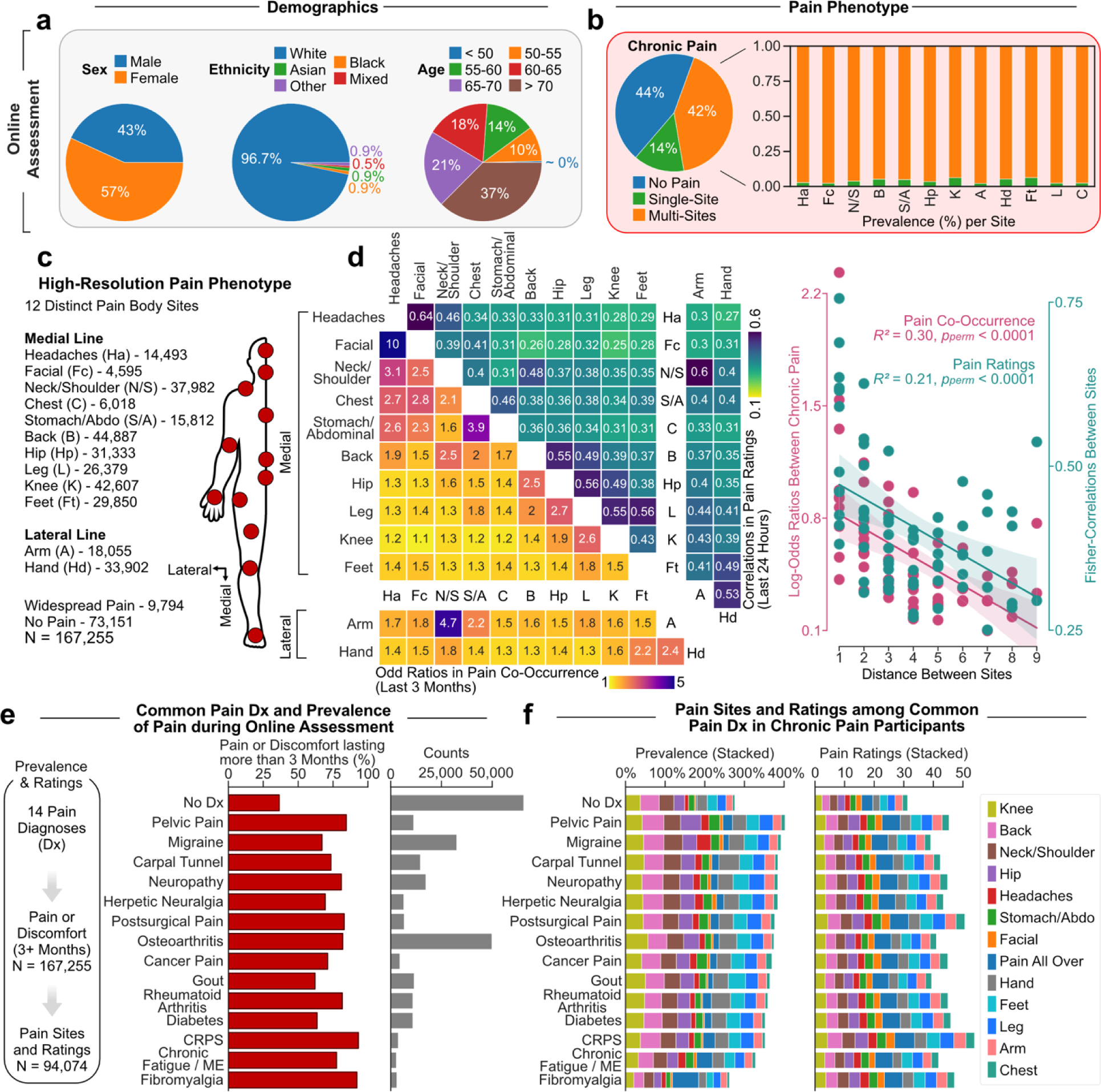
Online follow-up assessment of the experience of pain. **a.** Demographics of participants across sex, ethnicity, and age. **b.** Pain reported in the past 3 months (being chronic, > 3 months) for single and multi-site pain. **c.** High-resolution representation of anatomical body map sites and counts across a total of 13 sites: 10 along the medial line, 2 along the lateral line (shoulder to arm-hand) and 1 not localized (widespread). **d.** Cross-sectional analysis of co-existing pain and pain ratings. Odd ratios of co-occurrence between sites in the past 3 months (left diagonal, yellow) and Pearson’s correlation between pain ratings interdependence in the last 24 hours (right diagonal; green). Both the log-normalized odds ratio of pain sites and fisher-normalized odd ratios occurring in the past 3 months and fisher-normalized correlations were negatively associated with their distance. Significance was determined using 10,000 permutations tests. **e-f.** A total of 14 common chronic pain conditions diagnoses were included. **e.** Counts of diagnoses across the entire online assessment and the prevalence of those reporting pain or discomfort in the past 3 months. No Dx includes those without any of the 14 diagnoses. **f.** Pain prevalence and mean pain ratings (10 = as bad as you can imagine) across each diagnosis stacked across sites. Abbreviations: Headache (Ha); Facial (Fc); Neck or shoulder (N/S); Chest (C); Stomach or abdominal (S/A); Back (B); Hip (Hp); Leg (L); Knee (K); Feet (Ft); Arm (A); Hand (Hd).

**Extended Fig. 2.**
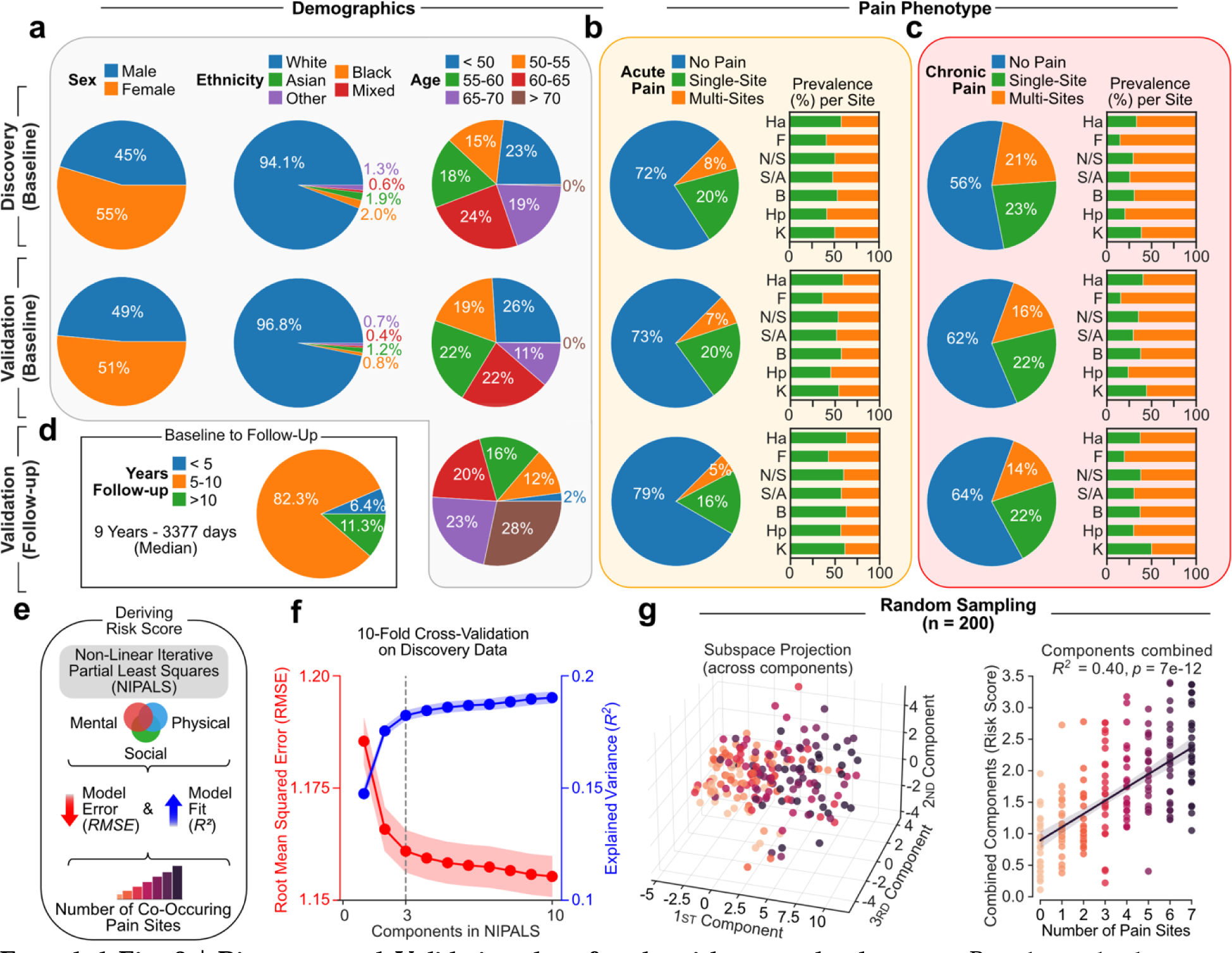
Discovery and Validation data for the risk score development. Pie charts displaying **a.** demographics, **b.** acute (≤ 3 months) and **c.** chronic (> 3 months) pain phenotype for the discovery data the model is trained on and the validation data the model is tested on, at baseline and follow-up. **d.** Years between baseline and follow-up visit in the validation data (9 years median). **e.** Schematic on using NIPALS to predict co-existing pain from biopsychosocial features. **f.** Model specification based on 10-fold cross validation by minimizing the root mean square (RMSE) and maximizing the explained variance (R^2^) average across 10-folds. Following the scree plot (elbow rule) criterion and to minimize overfit, 3 components were selected**. g.** Random stratified sampling of 200 participants projected across the 3 components separately and combined used as our risk score. Headache (Ha); Facial (F); Neck or shoulder (N/S); Stomach or abdominal (S/A); Back (B); Hip (Hp); Knee (K).

**Extended Data Fig 3.**
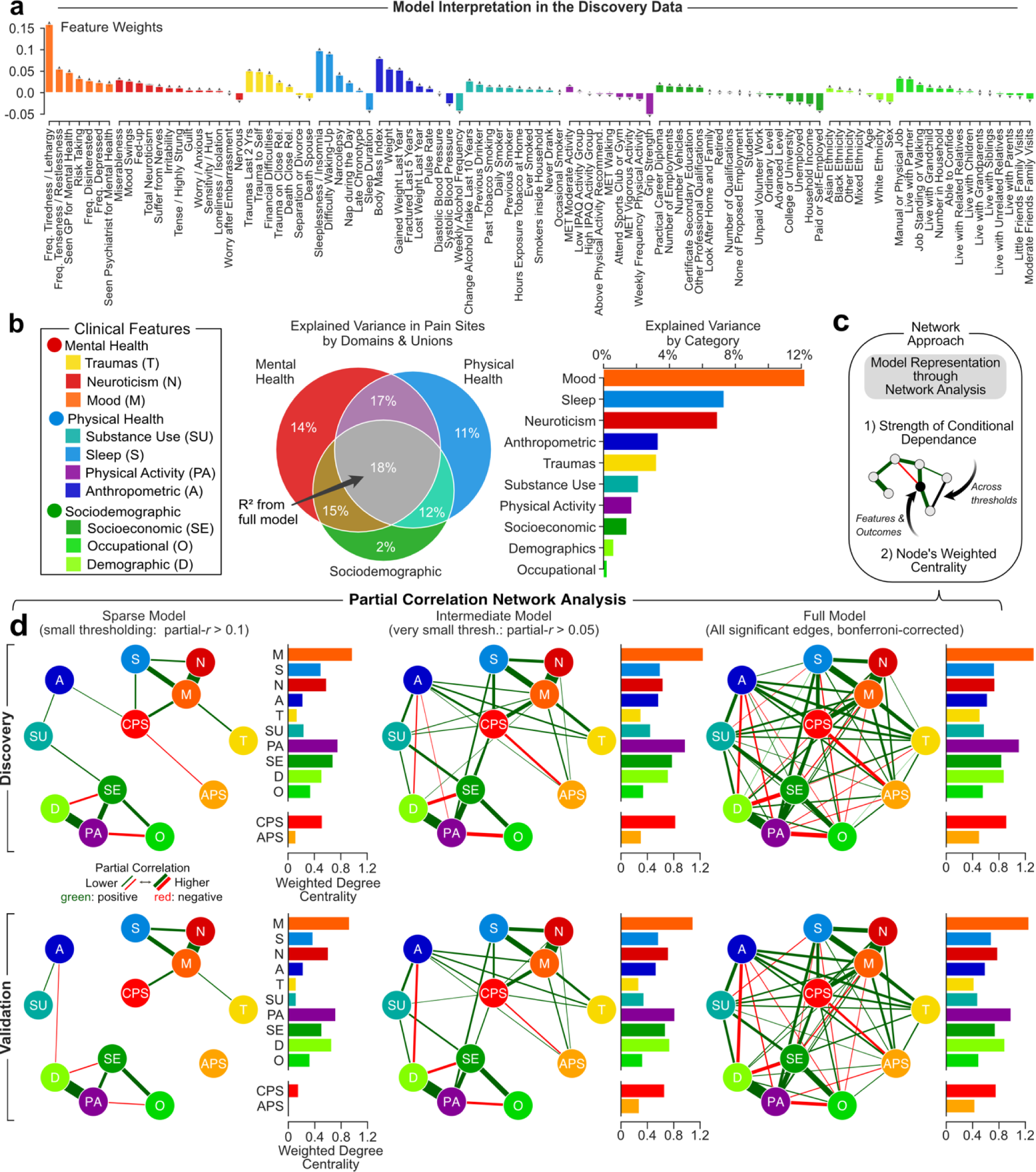
Detailed model interpretation in the discovery data. **a.** Model features weights to project to the risk score. **b.** Venn diagram and bar graphs of the explained variance across domains and their unions and categories in the discovery data. **c.** Schematic of a network approach to integrate the risk model’s categories. Edges and nodes were evaluated using strength of partial correlation and weighted node centrality. **d.** Partial correlation networks analysis across three levels: sparse (absolute partial correlation above 0.1), intermediate (above 0.05), and full (all edges) across the discovery data (upper row) and validation data (lower row). Abbreviations: Trauma (T); Neuroticism (N); Mood (M); Substance Use (SU); Sleep (S); Physical Activity (PA); Anthropometric (A); Socioeconomic (SE); Occupational (O); Demographic (D).

**Extended Fig. 4.**
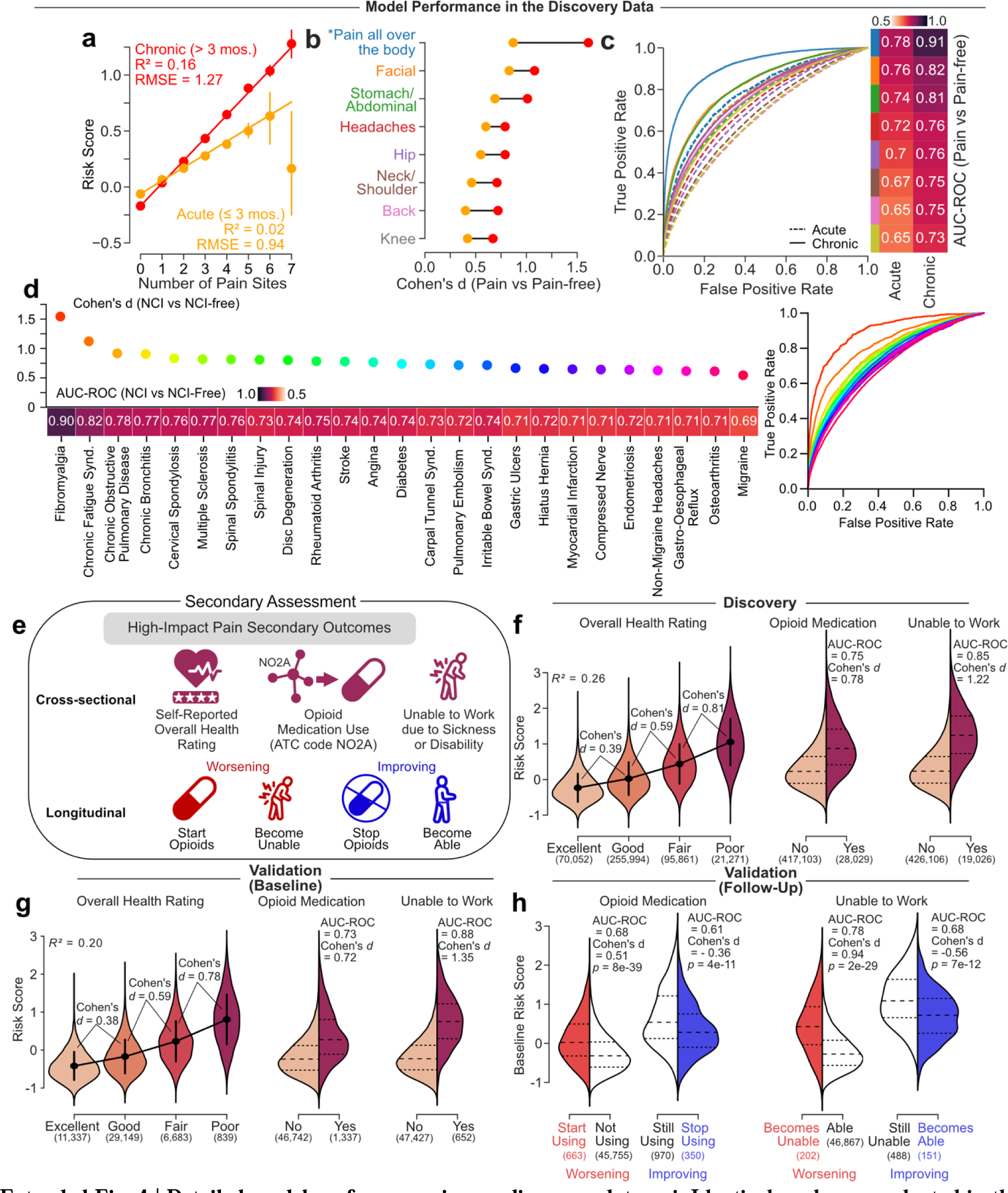
Detailed model performance in our discovery data. **a-i.** Identical analyses conducted in the original discovery from which the model was derived on (see Fig. 2. **e-h.** Evaluating the model performance in predicting secondary outcomes associated with high-impact chronic pain. **e.** Schematic of three pain-related secondary outcomes used to assess the clinical impact of pain. **f-g.** Cross-sectional performance of the secondary outcomes. Cohen’s d effect sizes and explained variance (R^2^, on the left) were used across self-reported ratings of overall health ratings while Cohen’s d and AUC-ROC discriminations were used for opioid medication use and inability to work due to sickness or disability in **f.** the discovery data and **g.** the validation data. **h.** Longitudinal prognosis two secondary outcomes from the risk score at 9-years follow-up. Cohen’s d and AUC-ROC were measure in the worsening at follow-up (left in red) and improvement (right in blue). Non-Cancer Illness (NCI); Area Under the Curve (AUC); Receiver Operating Characteristic (ROC). Sample sizes are included in parenthesis.

**Extended Fig. 5.**
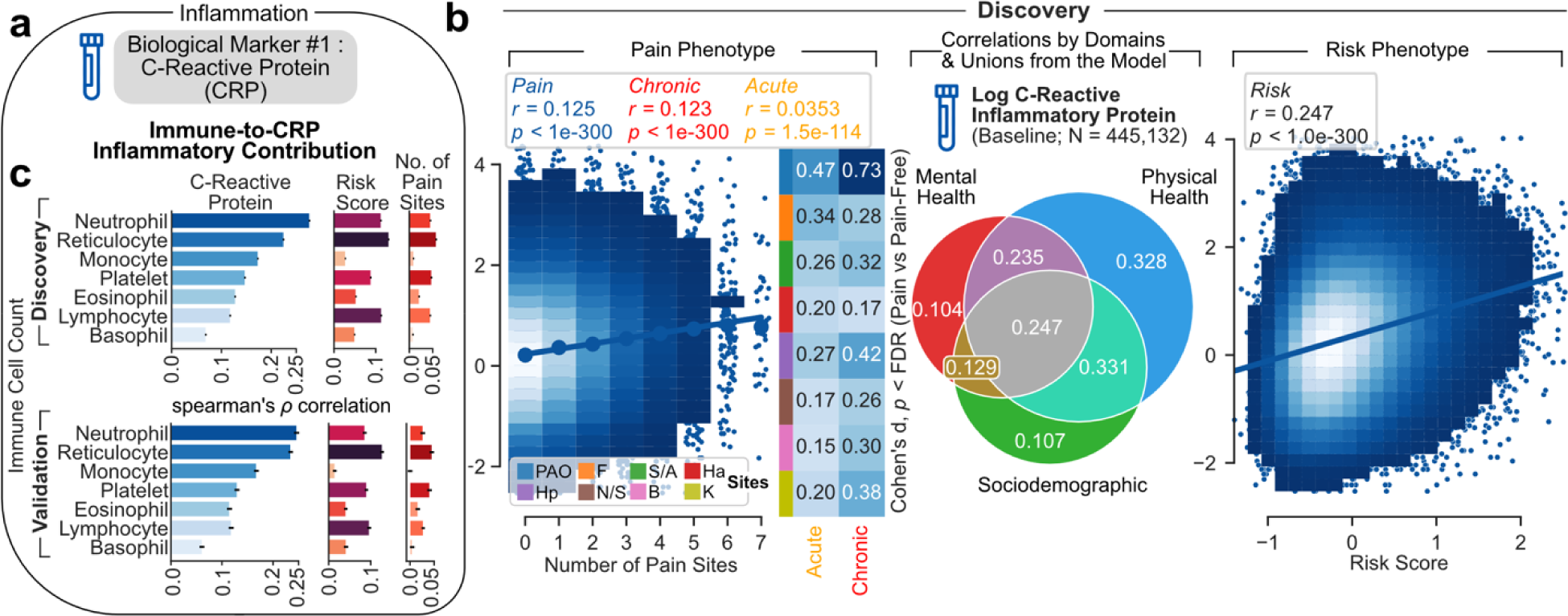
C-Reactive Protein (CRP) as main inflammatory marker. **a.** CRP was chosen as main inflammatory marker. **b**. The association between CRP (log-transformed for parametric estimations) and the number of pain sites and our risk score. The Venn diagram shows the correlation between the CRP with respect with the 3 domains. The Cohen’s d effect sizes are presented for each pain site compared to pain-free individuals. Comparisons were FDR-corrected (q = 0.05). **c.** Spearman’s rank association between immune cell counts and CRP, our risk score, and the number of pain sites in discovery and validation set. Errors bars were estimated using 1,000 resampling.

**Extended Fig. 6.**
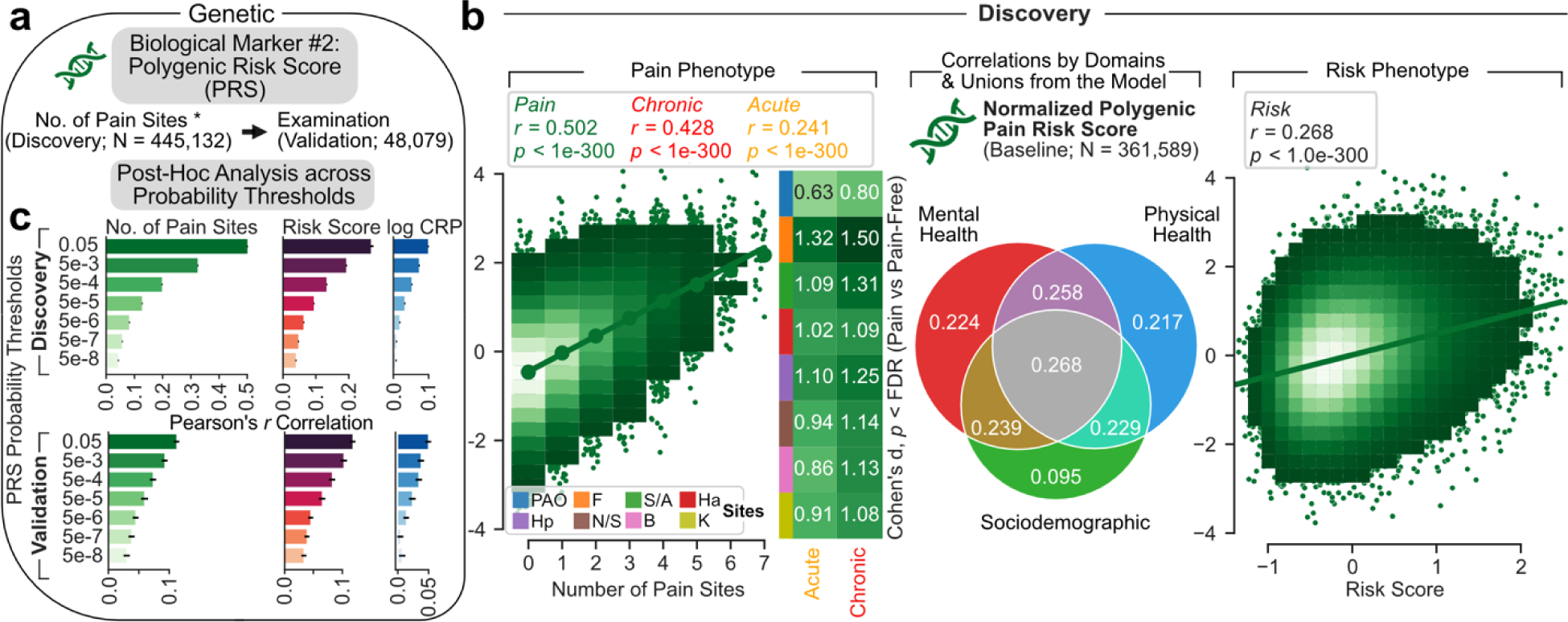
Polygenic Risk Score (PRS) for Genetic Risk of Pain. **a.** Genome-wide association study was done on the discovery set to derive a Polygenic Risk Score (PRS) on the number of pain sites, done across an array of thresholds. The least stringent threshold was taken to maximize prediction. **b**. The association between the most normalized PRS and the number of pain sites and our risk score. The Venn diagram shows the correlation between the PRS with respect with the 3 domains. The Cohen’s d effect sizes are presented for each pain site compared to pain-free individuals. Comparisons were FDR-corrected (q = 0.05). **c.** Pearson’s r association between each PRS and the number of pain sites, our risk score and log-transformed C-Reactive Protein (CRP) in the discovery and validation set. Errors bars were estimated using 1,000 bootstrap resampling.

**Extended Fig. 7.**
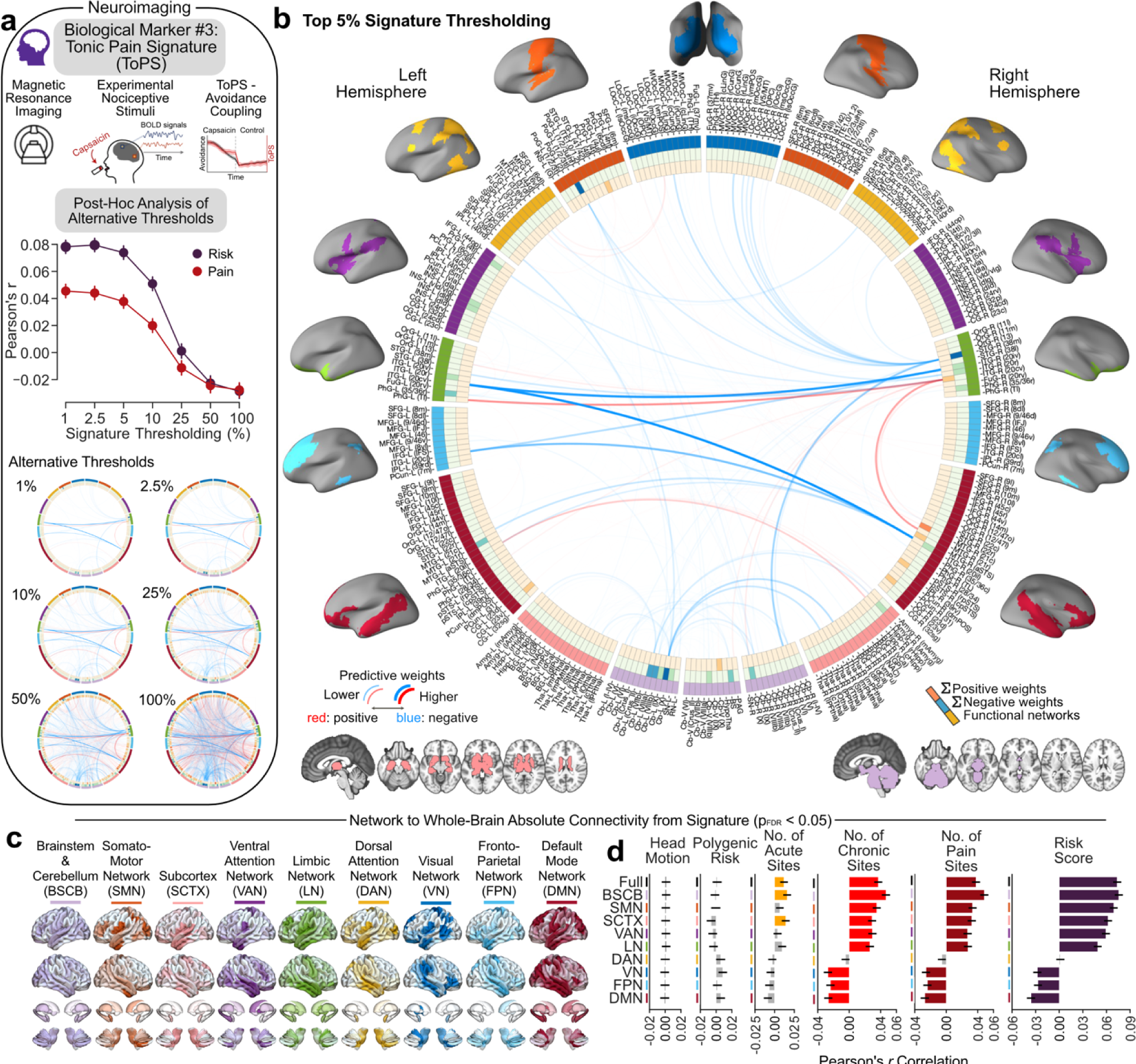
Tonic Pain Signature (ToPS) for the neuroimaging marker. **a.** Tonic Pain Signature (ToPS) was used to capture blood oxygenated-dependant level fluctuation following a capsaicin-induced sustained pain. Top 5% weights of the signature were used. Alternatives thresholds and the top 5% thresholds shown in **a.** and **b.** respectively. **c.** Connectivity patterns from the top 5% signature were extracted from each of the major networks. **d.** The association with head motion, our selected polygenic risk score, the number of acute, chronic, and combined pain sites as well as our risk score with each network. Comparisons were FDR-corrected (q = 0.05, non-significance shown in grey). Errors bars were estimated from 10,000 bootstrap resampling. Abbreviations: Brainstem and Cerebellum (BSCB); Somatosensory Network (SMN); Subcortex (SCTX); Ventral attention Network (VAN); Limbic Network (LN); Dorsal Attention Network (DAN); Visual Network (VN); Fronto-Parietal Network (FPN); Default Mode Network (DMN).

**Extended Fig. 8.**
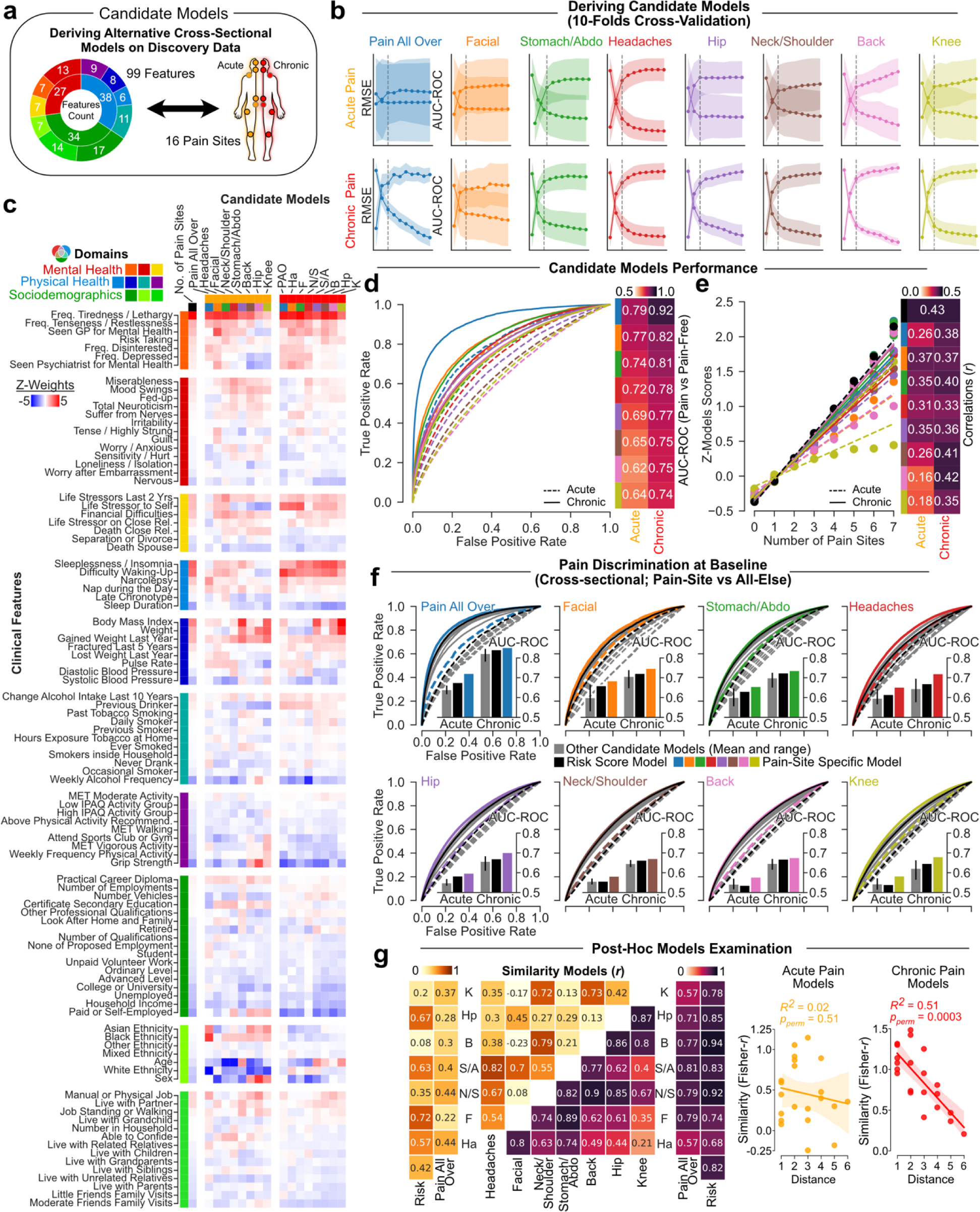
Deriving candidate models for chronic and acute pain conditions. **a.** A schematic describing the 99 features to derive a total of 16 site-specific candidate models cross-sectionally in the discovery set**. b.** 10-Fold cross-validation was used to estimate the root mean square (RMSE) and explained variance (R^2^). The same number of components were used to ensure comparability between derived models using NIPALS. **c.** Weights used for each model (normalized to allow comparison) grouped across categories and domains. **d.** Candidate models’ capacities to discriminate between the pain sites they were trained on from pain-free individuals are shown using the Area Under the Receiver Operator Characteristic Curve (AUC-ROC). **e.** The risk score derived from each candidate model correlated with number of co-existing pain sites. **d.** Cross-sectional discrimination for each pain site in acute (dashed line) and chronic (full line) pain conditions against the rest of the training cohort (i.e., pain-free and other pain sites) using the model specific to the site (in color), to the number of pain sites (black), and to other candidate models trained on a different pain site (grey). **f.** Post-hoc analyses show that similarities between the weights of the different models are associated with the distance between the sites for chronic pain conditions. Root mean squared error (RMSE); Area Under the Curve (AUC); Receiver Operating Characteristic (ROC); Headache (Ha); Facial (F); Neck or shoulder (N/S); Stomach or abdominal (S/A); Back (B); Hip (Hp); Knee (K).

